# Genome-wide gene-air pollution interaction analysis of lung function in 300,000 individuals

**DOI:** 10.1101/2021.06.03.21256376

**Authors:** Carl A. Melbourne, A. Mesut Erzurumluoglu, Nick Shrine, Jing Chen, Martin D. Tobin, Anna Hansell, Louise V. Wain

## Abstract

Impaired lung function is predictive of mortality and is a key component in the diagnosis of chronic obstructive pulmonary disease. Lung function has a strong genetic component but is also affected by environmental factors such as increased exposure to air pollution. How genetic factors and air pollution interact to affect lung function is however less understood.

We conducted a genome-wide gene-air pollution interaction analysis of spirometry measures with three measures of air pollution at home address: particulate matter (PM_2.5_ & PM_10_) and nitrogen dioxide (NO_2_), in approximately 300,000 unrelated European individuals from UK Biobank. We explored air pollution interactions with previously identified lung function signals and determined their combined interaction effect using a polygenic risk score (PRS).

We identified seven genome-wide interaction signals (*P* < 5 × 10^−8^), and a further ten suggestive interaction signals (*P* < 5 × 10^−7^). We found statistical evidence of interaction with PM_2.5_ for previous lung function signal, rs10841302, near *AEBP2*, suggesting increased susceptibility of FEV_1_/FVC to PM_2.5_, as copies of the G allele increased (interaction beta: −0.073 percentage points, 95%CI: −0.105, −0.041). There was no observed interaction between air pollutants and the weighted genetic risk score.

We carried out the largest genome-wide gene-air pollution interaction study of lung function and identified effects of clinically relevant size and significance. We observed up to 440ml lower lung function for certain genotypes associated with mean levels of outdoor air pollution at baseline, which is approximately equivalent to nine years of normal loss of lung function.

## Introduction

Impaired lung function is predictive of mortality and is a key component in the diagnosis of chronic obstructive pulmonary disease (COPD). Smoking is the biggest risk factor for COPD, which is thought to have caused as many as 2.9 million deaths worldwide in 2016 (1) although other sources of indoor air pollution are also associated with COPD risk (2, 3). Furthermore, increased exposure to air pollution is associated with lower lung function (4).

Lung function and COPD risk is also influenced by genetic factors and we and others have discovered over 300 genetic association signals for COPD risk and/or lung function measures (5, 6). Combining these signals into a single polygenic risk score, we have shown that individuals in the highest decile of genetic risk have an almost 5-fold increased risk of COPD compared to those in the lowest decile. However, collectively, these variants only explain up to around 13% of the heritability of lung function.

We hypothesised that there could be interactions between genetic variants and air pollution measures which affect COPD risk and lung function. Detection of such effects could enable identification of high-risk subgroups of the population and provide new biological insight into the mechanisms whereby air pollution affects respiratory health.

To test this hypothesis, we carried out the largest genome-wide gene-air pollution interaction study of lung function in ~300,000 individuals from UK Biobank, using particulate matter (PM) and nitrogen dioxide (NO_2_) concentrations as measures of air pollution exposure.

## Methods and Materials

### Selection of individuals with lung function data

We selected unrelated European individuals from UK Biobank as previously described (6). In summary, we selected individuals that had complete lung function data and passed our previously outlined quality control filters for forced expiratory volume in 1 second (FEV_1_), forced vital capacity (FVC) and the ratio (FEV_1_/FVC). From this we then selected a subsample of unrelated individuals of genetically determined European ancestry (KING kinship coefficient < 0.0884 corresponding to below 2^nd^ degree kinship (7)). All individuals had complete data for sex, age, height and ever smoking status (ever vs never).

### Air pollution data

Air pollution concentrations at place of residence of UK Biobank participants at recruitment were estimated using European Study of Cohorts and Air Pollution Effects (ESCAPE) land use regression models (8, 9). In these analyses, we explored associations with fine particles with average diameter < 2.5 µm (PM_2.5_), particulate matter with average aerodynamic diameter < 10 µm (PM_10_) and annual average concentrations of nitrogen dioxide (NO_2_).

For the particulate measures, models were not robust more than 400km from Greater London, so analyses did not include participants from northern England and Scotland. ESCAPE NO_2_ variables were available UK-wide.

### Genome-wide interaction analysis

FEV_1_, FVC and FEV_1_/FVC were adjusted for sex, age, age^2^, height and ever smoking. Residuals were then inverse normal transformed. Air pollution measures PM_2.5_ and PM_10_ were transformed into standard z-scores to avoid collinearity issues between the air pollution and interaction terms (observed due to small measurement variances). Air pollution measure NO_2_ was analysed untransformed.

Individuals were genotyped using the Affymetrix Axiom UK BiLEVE and Affymetrix Axiom UK Biobank arrays (10) with imputation undertaken using the Haplotype Reference Consortium (HRC) (11)and combined UK10K + 1000 genomes (12) reference panels. Multiallelic variants were removed and variants imputed with low confidence were excluded (imputation quality r^2^ < 0.5 for all SNPs and r^2^ < 0.8 for rare SNPs with minor allele frequency (MAF) < 1%). Variants with MAF less than 0.5% were removed.

Each transformed lung function trait was used as the outcome in a multiple regression model which included the first 15 principal component terms for ancestry, genotyping array, SNP term (using an additive genetic model), air pollution variable and an interaction term for the interaction between SNP and air pollution:

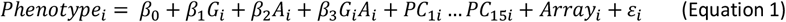

where *G*_*i*_ is the genotype for individual *i, A*_*i*_ is the air pollution value, *PC*_1*i*_ … *PC*_15*i*_ represent principal component values and *Array*_*i*_ is the genotype array value (coded 0 and 1 for UK Biobank array and UK BiLEVE array respectively). The p-value returned for the *β*_3_ estimate corresponds to the interaction effect between SNP and air pollution value (*G*_*i*_*A*_*i*_). Multiple regression was performed using PLINK2 (13).

### Signal selection and fine-mapping

To define association signals and their sentinel variants, all variants were ranked by p-value and the SNP with the lowest p-value was selected as the first signal sentinel. All SNPs +/−1 megabase (Mb) either side of this first sentinel were then excluded and the process repeated for the next most significant SNP until all 2Mb regions containing a sentinel SNP with *P* < 5 × 10^−8^ had been identified (genome-wide signals). The process was repeated to define a set of signals with sentinel SNPs at threshold of *P* < 5 × 10^−7^ (suggestive signals). Conditional analysis was used to identify additional independent genome-wide and suggestive signals by including the sentinel interaction term in the model, re-analysing all SNPs within each 2Mb region and determining whether any SNPs remained below the pre-specified threshold. Region plots for each signal were created using LocusZoom (14).

To aid the interpretation of interaction effects for genome-wide significant interaction signals, we presented the association between lung function trait and air pollution variable stratified by genotype group. To do this, dosages were converted to direct genotype calls by rounding to the nearest genotype group.

Using a Bayesian method (15) we fine-mapped each signal to a credible set of SNPs (the set of SNPs 95% likely to contain the causal SNP, under the assumption that the causal SNP was analysed).

### Identification of putative causal genes

Credible set SNPs including the sentinel SNP were annotated using Annovar (16) to identify coding variants with a putative functional effect (for example, missense). To identify whether any of the signals were independently associated with gene expression, we searched the GTEx (17) and blood eQTLgen (18)eQTL catalogues. To identify a potential shared causal variant between the SNP-air pollution interaction signals and the eQTL gene expression signals, colocalisation was undertaken using COLOC (19) where full summary data was available in GTEx and eQTLgen databases (20). An observed probability > 0.8 for a shared causal variant was used as the threshold to conclude colocalisation of SNP-air pollution and gene expression signals. We queried the sentinel SNPs in Open Target Genetics (21) for eQTL associations (which in addition to GTEx includes a further 14 consortia with eQTL expression association results) and to identify associations with protein expression (pQTL) and overlap with regions known to interact with gene promoters (promotor capture HiC).

### Association with other phenotypes

The SNP with the highest posterior probability for causality in each credible set was queried in PhenoScanner (22) and Open Targets Genetics (21) resources to identify shared associations with other phenotypes at a threshold of *P* < 1 × 10^−3^.

### Functional enrichment

To identify whether there was enrichment of SNP-air pollution interaction signals within regulatory regions of the genome (for example, DNase Hypersensitivity Sites) in specific cell or tissue types we used GARFIELD (23). The software determines whether signals are enriched for DNase I hypersensitive sites across 55 tissues (with an adjusted significant enrichment threshold for 540 effective annotations of P < 9.26 × 10^−5^). We investigated the functional impact of SNPs (potential chromatin effects) which were highly probable to be the drivers of each signal (i.e. SNPs with posterior probability > 0.9 in credible sets) using DeepSEA (24). To define a significant functional impact we used an E-value < 0.05 (the proportion of 1000 Genomes SNPs predicted to have a higher magnitude for chromatin effect compared to the chosen SNP being investigated) and an absolute probability difference > 0.1 between alternative and reference allele (the threshold defined for ‘high confidence’).

### Effects of Socio-Economic Status

Socio-economic status (SES) of an individual is a plausible moderator of lung function, with observed modification of air pollution effects (4), however adjusting for SES in our analyses would have led to a reduction of approximately 13% in the discovery sample size due to missing data. We accounted for any effects of SES on genome-wide interaction signals in two ways. Firstly, we undertook a sensitivity analysis for the top signals adjusting for educational status and income status using a complete-case analysis (after inverse normalisation of lung function traits). Secondly, we present interaction effects for genome-wide signals across categorised groups for income and educational status to visualise any difference in effect (akin to a three-way interaction between SNP, air pollutant and education/income). Income status was categorised using the definition in UK Biobank of “less than £18,000”, “£18,000 to £30,999”, “£31,000 to £51,999”, “£52,000 to 100,000” and “Greater than 100,000”. Educational status was dichotomised as “lower vocational qualification or less” vs “higher vocational qualification or more”, grouping A-level (2), O-level (3), CSEs (4), and “None of the above” (−7) under “low education”, and College/University (1), NVQ (5) and Other professional qualifications (6) under “high education”. Individuals who selected “Do not know” (−1), “Prefer not to answer” (−3) or have missing data were excluded from subsequent analyses.

### Previously reported lung function and COPD association signals

We performed a look-up in the genome-wide gene-air pollution interaction analyses (for all three air pollution measures and all three lung function measures), for the 304 signals previously reported for association with lung function and COPD (279 lung function signals from Shrine et al. 2019 (6) and 25 signals from Sakornsakolpat et al. 2019 (5)). As these independent signals have *a priori* evidence for association with lung function or COPD, we applied a Bonferroni corrected threshold for 304 tests to define a significant air pollution interaction effect (*P* < 1.6 × 10^−4^). As before, to aid interpretation of the interaction effect for any statistically significant signal, we present the association between lung function trait and air pollution stratified by genotype group.

### Weighted genetic risk score interaction analysis

We used a weighted polygenic risk score (PRS) to explore whether the combined effect of previously reported lung function signals showed an interaction with air pollution measures (i.e. whether the phenotypic effects of the SNPs were modified by exposure to air pollution). Each individual’s score was calculated using the effect sizes of the 279 SNPs reported in Shrine et al. 2019 (6) on FEV_1_/FVC (using the lung function reducing allele as the coded allele). Multiple regression was performed using the same model above, using the weighted polygenic risk score in place of the genotype. As all three lung function traits are correlated, interaction terms (i.e. wGRS x Air pollution measure) with *P* < 0.05 were defined as statistically significant.

### Antioxidant genes and their interaction with air pollution

Genetic variation within antioxidant genes may contribute to susceptibility of adverse effects of air pollution on respiratory health (25). We have provided look-ups for the most commonly evaluated antioxidant genes (for which a SNP was reported) and for SNPs evaluated in previous antioxidant-gene-air pollution interaction studies, both of which are reviewed in Fuertes et al. (25). A Bonferroni adjusted threshold of P < 3.85 × 10^−3^ (for 13 variants) was used to determine statistical significance.

## Results

### Genome-wide interaction analysis

Genome-wide interaction analysis was undertaken in 277,597 European individuals from UK Biobank for air pollution variables PM_10_/PM_2.5,_ (**Supplementary table 1**) and a total of 10,848,082 SNPs (**Supplementary figure 1**). For the NO_2_ analysis, there were 299,015 European individuals and 10,846,777 SNPs. Manhattan plots are presented in **Figure 1** and QQ plots in **supplementary figure 2**.

**Figure 1.**
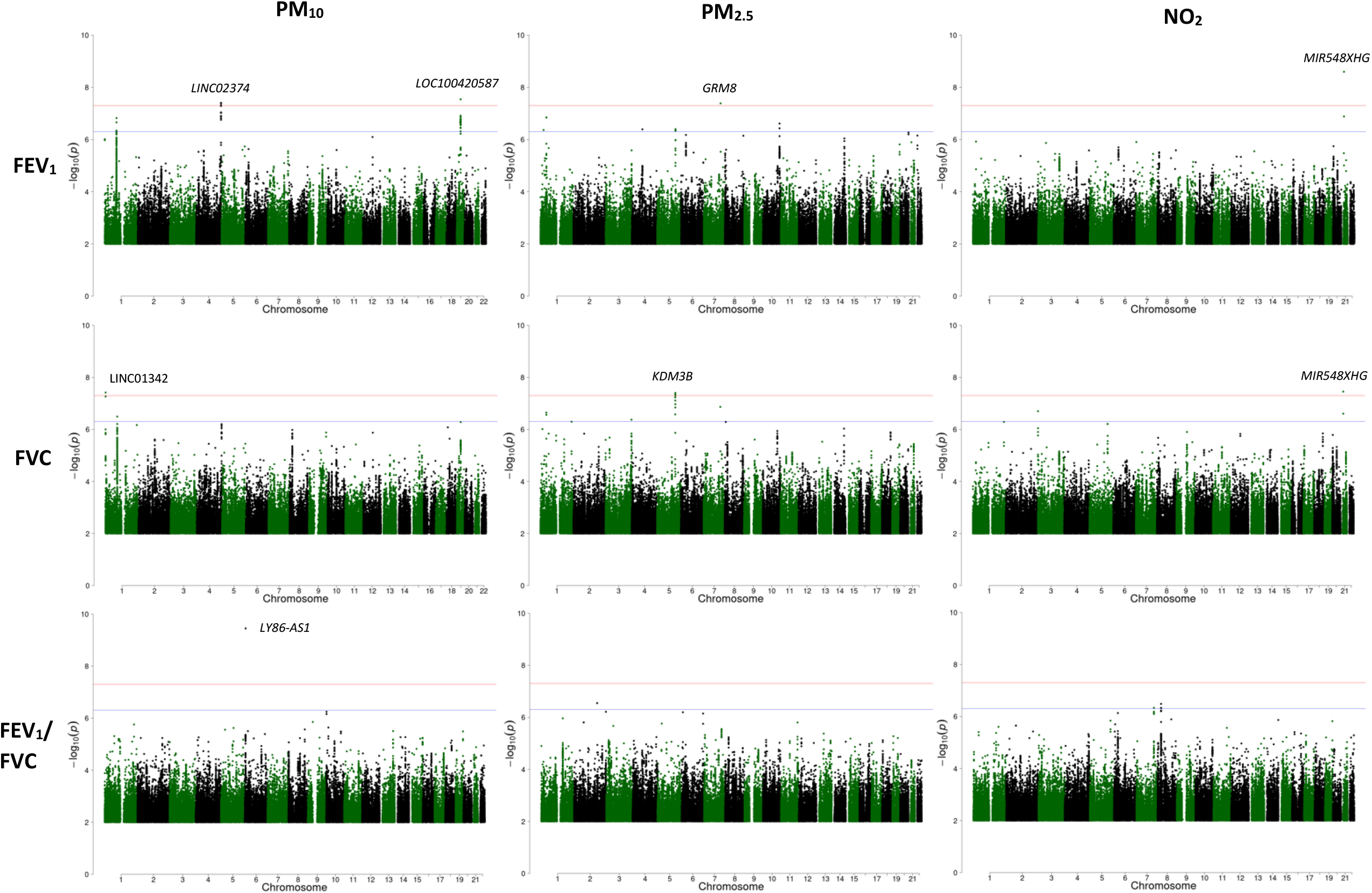
Manhattan plots for the gene-air pollution interaction GWAS. The red line represents a p-value threshold of 5 × 10^−8^. The blue line represents a p-value threshold of 5 × 10^−7^. Each genome-wide signal is annotated by nearest gene.

We identified seven signals with an interaction effect reaching genome-wide statistical significance (*P* < 5 × 10^−8^) for at least one lung function trait and air pollution variable (**Table 1, Supplementary table 2 and Supplementary figure 3**). Four signals were identified for an interaction with PM_10_. These included two for FEV_1_ (in 4q35.2 [near *LINC02374*] and in 19q12 [near *LOC100420587*]), one for FVC (in 1p36.33 [near *LINC01342*]) and one for FEV_1_/FVC (in 6p25.1 [in *LY86-AS1*]). Two signals were identified for an interaction with PM_2.5_; one for FEV_1_ (in 7q31.33 [near *GRM8*]) and one for FVC (in 5q31.2 [in *KDM3B*]. One signal was identified for air pollutant NO_2_ for both lung function traits FEV_1_ and FVC (in 21q21.1 [near *MIR548XHG*]). Of the seven identified SNPs, three were common (MAF > 5%) two were low frequency, (1% < MAF < 5%) and two were rare (MAF < 1%). Conditional analysis did not identify any additional signals in each region.

**Table 1.** Seven identified genome-wide gene-air pollution interaction signals, CAF = Coded Allele Frequency, INFO – imputation quality, AP = Air Pollutant, LF = Lung Function, BP = Base Position. A negative BETA (interaction effect) suggests a more deleterious effect on lung function per unit increase of air pollutant as the coded allele increases. A positive BETA (interaction effect) suggests a more protective effect. Interaction effect is per unit increase in air pollutant NO_2_ (1 μg/m^3^) and per standard deviation increase for air pollution variables PM_10_ and PM_2.5_ as the coded allele increases. Lung function effects are the product of the BETA value and the observed standard deviation of the lung function trait within the analysed sample.

| LF trait | AP | SNP | CHR | BP | Coded | Non Coded | CAF | INFO | BETA | SE | LF effect (ml for FEV<sub>1</sub>, FVC or % points for FEV<sub>1</sub>/FVC) | P | Locus |
| --- | --- | --- | --- | --- | --- | --- | --- | --- | --- | --- | --- | --- | --- |
| FVC | PM<sub>10</sub> | rs74048016 | 1 | 1068280 | C | G | 0.98 | 0.97 | -0.053 | 0.010 | -50.8 | 3.83×10<sup>-8</sup> | C1orf159 (dist=16811), LINC01342 (dist=4117) |
| FEV<sub>1</sub> | PM<sub>10</sub> | rs28666788 | 4 | 188078645 | G | A | 0.096 | 0.99 | -0.025 | 0.005 | -18.9 | 3.92×10<sup>-8</sup> | FAT1 (dist=433635), LINC02374 (dist=44495) |
| FVC | PM<sub>2.5</sub> | rs192415220 | 5 | 137726002 | C | T | 0.006 | 0.94 | -0.100 | 0.018 | -95.9 | 3.96×10<sup>-8</sup> | KDM3B |
| FEV<sub>1</sub>/FVC | PM<sub>10</sub> | rs137914543 | 6 | 6414006 | G | GTCTC | 0.06 | 0.83 | -0.038 | 0.006 | -0.24 | 3.55×10<sup>-10</sup> | LY86-AS1 |
| FEV<sub>1</sub> | PM<sub>2.5</sub> | rs138235384 | 7 | 125969169 | C | T | 0.994 | 0.90 | -0.097 | 0.018 | -73.3 | 4.10×10<sup>-8</sup> | LOC101928283 (dist=949794), GRM8 (dist=109483) |
| FEV<sub>1</sub> | PM<sub>10</sub> | rs762101031 | 19 | 29112275 | CAAT | C | 0.96 | 0.79 | -0.046 | 0.008 | -34.8 | 2.87×10<sup>-8</sup> | LOC100420587 |
| FEV<sub>1</sub> | NO<sub>2</sub> | rs2825255 | 21 | 20362376 | T | C | 0.83 | 1.00 | -0.003 | 0.001 | -0.23 | 2.53×10<sup>-9</sup> | MIR548XHG (dist=230246), LINC01683 (dist=903217) |
| FVC |  |  |  |  |  |  |  |  | -0.003 | 0.001 | -0.29 | 3.52×10<sup>-8</sup> |  |

To aid with the interpretation of statistically significant interaction effects, we have presented the association between air pollution and lung function stratified by genotype group (number of copies of coded allele) for each of the seven genome-wide interaction signals (**Figure 2)** and interaction plots of predicted lung function against air pollution for each genotype group (**Supplementary figure 4)**. In some instances, statistically significant association between lung function and air pollutant is observed in all genotype groups. For others, the association only reaches statistical significance for certain genotype groups.

**Figure 2.**
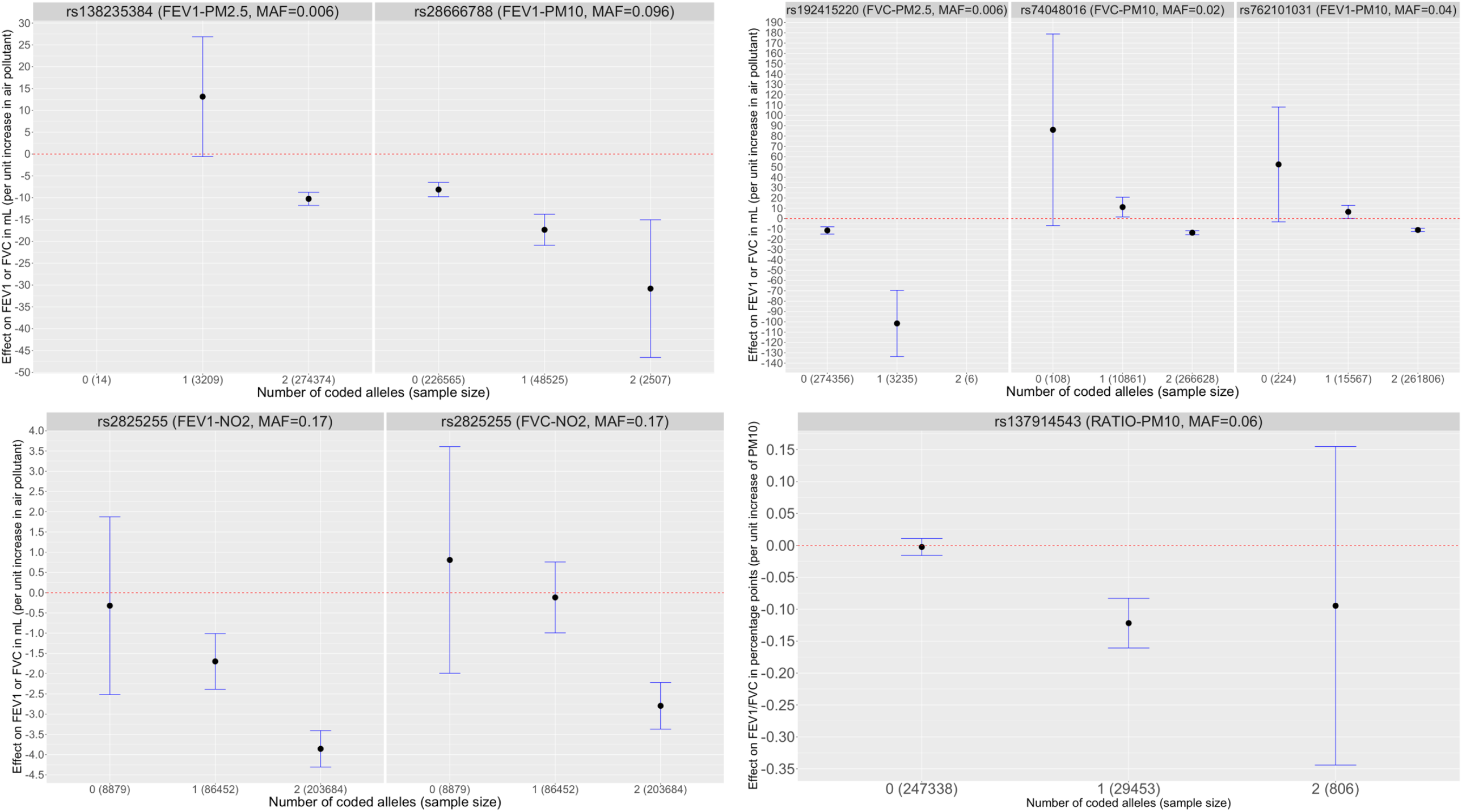
Association between lung function trait and air pollutant (effect size and confidence intervals) for the seven genome-wide signals. Note: For SNPs rs138235384 and rs192415220 the effect size for 0 copies and 2 copies of the effect allele respectively are not presented due to the low minor allele frequency and small sample size. Effect sizes will not be exactly consistent with Table 1 due to rounding error when converting from dosage to direct genotypes.

Signals were deemed suggestively statistically significant using the same signal selection procedure with a threshold of 5 × 10^−7^ (**Supplementary table 3 and Supplementary figure 5**). Region plots after conditional analysis suggested only one signal per 2Mb region. Ten suggestive signals were identified that were independent of the seven genome-wide significant signals, all were either intergenic or mapped to the intronic region of the mapped gene. Eight were represented by common SNPs, and two by low frequency SNPs.

### Credible sets and causal genes

To determine a causal gene for each signal (both genome-wide and suggestively significant), we used Bayesian fine mapping to define the 95% credible set of causal SNPs (assuming the causal SNP was included in the analysis, **Supplementary table 4**) and investigated whether credible set and sentinel SNPs were associated with changes in gene expression in GTEx, Blood eQTL and Open Target Genetics databases (**Supplementary table 5**). Genome-wide significant signal, rs74048016, whose C allele had a larger deleterious effect on lung function as the measurement of PM_10_ increased, was associated with decreased expression of *HES4* and increased expression of *C1orf159* and RP11-465B22.3 in blood. The signals did not colocalise with the known gene expression signals in this region (using the eQTLgen database (20)). Credible set SNPs for suggestive signals rs769937512, rs111552599, rs139556451, rs200460259 and rs10082259 were associated in various tissues for genes *AL445991*.*1, FRAS1, PNMA2*/*DPYSL, MUC4*/*MUC20* and *UROD* respectively (**Supplementary table 5**). These signals did not colocalise. There was no association with protein expression and no overlap with regions that had strong evidence for interaction with gene promoters.

### Association with other phenotypes

Sentinel SNPs were queried in PhenoScanner and Open Targets Genetics resources (**Supplementary table 6**). Five signals were found to be associated with at least one trait at *P* < 1 × 10^−3^, three genome-wide signals (rs28666788, rs192415220 and rs138235384) and two suggestive signals (rs10082259/rs6661026 and rs769937512). None of the associations reached genome-wide significance (*P* < 5 × 10^−8^). For the genome-wide signals rs28666788, rs192415220 and rs138235384 the strongest associations were with alcohol consumption, self-reported cervical polyps and sexual dysfunction respectively (at < 5 × 10^−6^).

### Functional enrichment

When looking for evidence of significant functional enrichment at DNase I hypersensitive sites (DHS) using GARFIELD, SNPs showing SNP-NO_2_ interaction effects on lung function phenotype FVC were enriched in various tissues including fetal lung, using a threshold of ***P*** < **5** × **10^−5^** to select contributing SNPs (**Supplementary figure 6**). No significant functional enrichment was observed when considering genome-wide statistically significant SNPs only (***P*** < **5** × **10^−6^**) or for any of the other eight combinations of lung function and air pollution measures. For the six SNPs which were highly probable to be causal drivers of their respective signals (posterior probability > 0.9), none showed any evidence of any chromatin effects using DeepSEA (**Supplementary table 7**).

### Effects of Socio-Economic Status

When adjusting for socio-economic status variables educational status and income status, sample sizes were reduced to 259,130 and 240,202 for the NO_2_ and PM_10_/PM_2.5_ analyses respectively. Effect sizes were largely consistent with the primary analysis with minimal reductions in effect size for rs74048016 and rs192415220 (**Supplementary table 8 and Supplementary figure 7**), suggesting that the interactions identified were not due to confounding by SES factors. Interaction effects were generally larger in magnitude (but not significantly due to overlapping confidence intervals) for those in the lower educational group (**Supplementary figure 8**). When stratifying by income group (**Supplementary figure 9**), overlapping confidence intervals again suggested no significant effect of income status on air pollution and lung function association across genotype groups. A slight inverse correlation between magnitude of interaction effect and income group was observed for rs2825255 for both lung function traits (higher income group, smaller interaction effect magnitude) with a positive correlation observed for rs762101031 (higher income group, larger interaction effect magnitude).

### Lung function associated signals

To determine whether any signals previously shown to be associated with lung function produced an interaction effect with air pollution variables, we performed a look up of the 304 variants (279 lung function signals from Shrine et al. (6) and 24 COPD signals from Sakornsakolpat et al. (5)) in our genome-wide analysis. Of the 304 signals, one signal, rs10841302, near *AEBP2*, for which the G allele is associated with lower values of FEV_1_/FVC, met a Bonferroni threshold of P < 1.6 × 10^−4^ for an interaction with PM_2.5_ for FEV_1_/FVC (interaction *β*: −0.0118; CI: −0.0170, −0.0066; interaction P=9.65×10^−6^) (**Supplementary table 9)**, suggesting a larger deleterious effect of PM_2.5_ on FEV_1_/FVC as copies of the G allele increased (**Figure 3**). This is equivalent to an FEV_1_/FVC effect of −0.073 percentage points (CI: −0.105, −0.041) per unit increase in PM_2.5_. The interaction can also be interpreted by air pollution and lung function association stratified by genotype group. For genotype groups CC, CG and GG for SNP rs10841302, a unit increase in PM_2.5_ (1 *μ*g/m^3^) resulted in a reduction of FEV_1_/FVC by 0.032 (95% CI: 0.026-0.038; P = 1.21 × 10^−22^), 0.034 (95% CI: 0.028-0.040; P = 5.52 × 10^−39^) and 0.056 (95% CI: 0.048-0.064; P = 2.43 × 10^−43^) standard deviations. This equates to direct FEV_1_/FVC effects of 0.19 (95% CI: 0.16-0.23), 0.20 (95% CI: 0.17-0.24) and 0.33 (95% CI: 0.29-0.38) percentage points respectively per 1 *μ*g/m^3^ of PM_2.5_.

**Figure 3.**
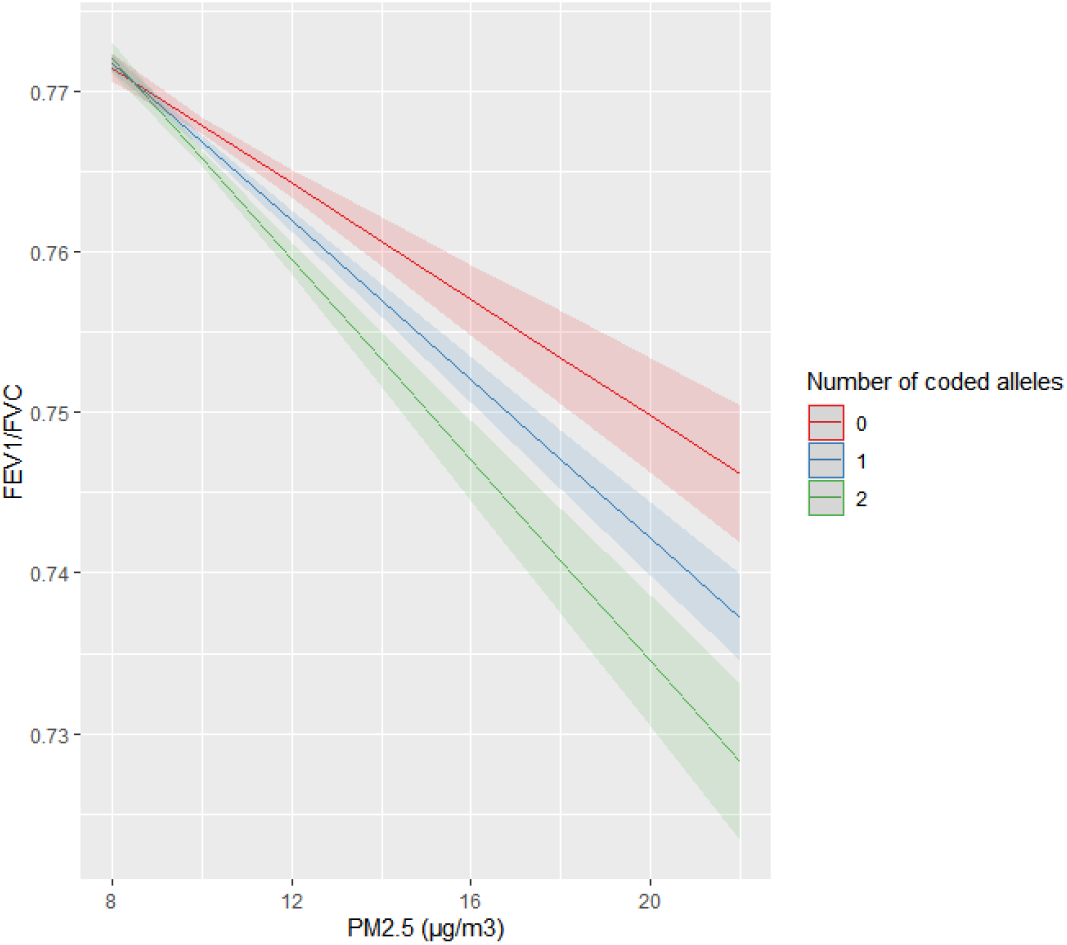
Interaction plot of FEV_1_/FVC predicted values against PM_2.5_ values across genotype groups (with coded allele G) for the previously identified lung function signal rs10841302

We tested the interaction between a weighted polygenic risk score for lung function (based on the effect sizes of 279 lung function signals reported in Shrine et al. (6)) and each air pollution measure on FEV_1_, FVC and FEV_1_/FVC (**Supplementary table 10**). None of the interaction effects were statistically significant (all P > 0.05).

### Antioxidant genes and their interaction with air pollution

We performed a look up of the 13 variants corresponding to seven commonly evaluated antioxidant genes and/or those analysed in previous studies of antioxidant gene-air pollution interaction analyses, as reviewed by Fuertes et al. (25) (**Supplementary table 11**). None of the SNPs reached the Bonferroni significant adjusted threshold used to determine statistical significance (P < 3.85 × 10^−3^). One SNP, rs1001179 in *CAT* approached this threshold (P = 0.009) for an interaction with NO_2_ for lung function phenotype FEV_1_/FVC.

## Discussion

We carried out the largest genome-wide gene-air pollution interaction study of lung function and identified seven genome-wide statistically significant signals, as well as identifying an interaction with air pollution for one previously identified lung function signal.

For the signals identified, ascribing the biological mechanisms proves a challenge and further biological studies of gene function for those implicated are needed. For genome-wide SNP rs74048016, as the number of copies of the coded allele increases the effect of air pollutant PM_10_ on FVC becomes more negative, suggesting that those with two copies of the effect allele are at increased susceptibility of air pollution effects. The coded allele is associated with decreased expression of *HES4* and increased expression of *C1orf159* in blood in Open Targets Genetics. The signals for genome-wide association and gene expression signals did not colocalise (there was insufficient evidence of a shared causal variant between the two analyses) in this genomic region (using data from eQTLgen). Expression of *HES4 (*hes family bHLH transcription factor 4) has been implicated in poor outcomes for patients with Triple Negative Breast Cancer (TNBC) (26) and both HES4 and *C1orf159* (chromosome 1 open reading frame 159) have been implicated via functional annotation (nearest gene) of other genome-wide significant loci for several traits and diseases, including peak expiratory flow (PEF) (27, 28). There is also evidence of colocalisation between gene expression and genome-wide analyses for these genes in certain tissues for height phenotypes (standing and sitting) (27, 28).

We identified a further ten signals (independent of the primary genome-wide signals) at suggestive statistical significance, which would be important to take forward in future replication analyses. Genes implicated include *PNMA2, DPYSL2* and *BNIP3L*, all via functional annotation of other genome-wide significant loci for height, and additionally for educational attainment phenotypes (29, 30). There was however no attenuation of suggestive signal rs139556451 (which implicated the aforementioned genes in our analysis) when re-analysing with adjustment for education and income status (in the subset for which this data was available). *BNIP3L* expression has also been linked with lung cancer (31). Additionally, gene *FRAS1* identified by eQTL associations for SNPs in the rs111552599 suggestive signal credible set has been implicated by other genome-wide signals for lung function, specifically for trait FEV_1_/FVC (6, 29) and mutations in *FRAS1* have been observed amongst individuals with Fraser syndrome, which can cause airway abnormalities (32, 33). *MUC4* (identified by credible set eQTL associations for rs200460259), which encodes airway mucins (34) is associated with severity of lung disease in cystic fibrosis (through functional annotation of another genome-wide signal) (35) and risk of lung cancer (association with variants in the gene) (36). We were however unable to determine whether the association signal for the genes described here were driven by the same causal variant as the interaction signal.

We identified an interaction effect between SNP rs10841302 (a previously identified lung function signal associated with FEV_1_/FVC) and PM_2.5_ for lung function trait FEV_1_/FVC. Previous work has shown that the rs10841302 G-allele is associated with a deleterious effect on FEV_1_/FVC. We found that this deleterious effect increased in magnitude as the exposure to PM_2.5_ increased. A causative gene for the association between rs10841302 and lung function has not been determined. The SNP is near *AEBP2* (AE Binding Protein 2), a transcriptional repressor with a possible contribution to histone methylation and the G allele is associated with increased expression of both *RP11-405A12*.*2* (in pancreas and subcutaneous adipose tissues) and *<u>RP11-664H17</u>*.*<u>1</u>* (in pancreas and tibial nerve tissues) in GTEx (17). There was no evidence of an interaction between air pollution measures and a combined effect from all previously identified lung function signals represented by a polygenic risk score.

A particular strength of this study is the discovery sample size available for the interaction analysis. Interactions are challenging to identify due to the requirement of much larger sample sizes than GWAS efforts exploring the marginal effects of genetic variants (37). This strength is however unfortunately a contributor to its biggest limitation, which is identifying suitable independent datasets of sufficient sample size with lung function data in European ancestry populations to replicate discovery interaction signals. We calculated that sample sizes to replicate three of our novel genome-wide interaction signals when considering the reported interaction effect, main genetic effect and air pollution variable effect (chosen from each MAF frequency group of common, low frequency and rare), signals rs28666788 (MAF = 10%), rs74048016 (MAF = 2%) and rs192415220 (MAF = 0.6%) would be ~72k, ~71k and ~66k respectively to detect the effect at 80% power. However, these sample sizes are indeed sensitive to any observed error in interaction effect estimates, such that when using lower and upper confidence interval effect estimates, sample sizes required could range from ~35k to ~194k.

The discovery of gene-air pollution interactions which affect lung function susceptibility is limited, likely due to the aforementioned difficulty in identifying suitable sample sizes to provide adequate power. Previous genome-wide interaction studies are either attributed to related phenotypes, such as asthma (38) or have focussed on candidate genes, such as those with a role in oxidative stress, where conclusions drawn are often inconsistent with respect to direction of effect or presence of interaction (39, 40). Previous studies of interactions between genes and smoking behaviour, the largest risk factor for poor lung function and COPD, have also been largely unsuccessful in identifying interaction signals. This has been of interest as not all smokers develop restrictive lung problems. Candidate gene-smoking interactions have been identified, however utilising small sample sizes with absence of replication (41–43) and none of the previously identified lung function signals produced an interaction with smoking behaviour (6). Genome-wide interaction analysis efforts have also been considered for lung function (44) however with little success, and although a recent study of gene-smoking interaction effects for COPD found a genome-wide significant interaction at *15q25*.*1* (45), this is likely driven by the strong association between this locus and smoking behaviour (46–48). There has however been some evidence of interaction between smoking behaviour and genetic risk scores, when combining the effects of SNPs associated with lung function (6, 49). To the best of our knowledge, no genome-wide significant smoking interaction signals for lung function have been identified, highlighting the impact of identifying novel genome-wide gene-air pollution interaction signals.

Should the interaction effects be replicated in future analyses, the magnitude of effects observed here suggest potential for clinically relevant impacts on those with certain genotypes. Results (**Table 1, Figure 2**) are expressed per unit pollutant. For context, average annual concentrations of PM_10_ in 2018 were 14.7 µg/m^3^ in 2018 at urban background air quality monitoring sites (likely to represent where most of the UK population live) (50). Corresponding concentrations for PM_2.5_ and NO_2_ was 10.0 µg/m^3^ and 20.1 µg/m^3^ respectively. Taking genome-wide signal rs28666788 as an example, (with coded allele G frequency of 0.096), effects on FEV_1_ per unit increase in PM_10_ were statistically significant for all genotype groups. For those with zero, one and two copies of the effect allele, lung function effects of approximately −7.5ml, −17.5ml and −30ml were observed per 1µg/m^3^ PM_10_ respectively (**figure 2**). Therefore, when subjected to the average concentrations of 14.7 µg/m^3^ of PM_10_, this equates to respective reductions of 112.5ml, 260ml and 440ml. Average declines in FEV_1_ per year could be up to 46ml for individuals aged 30 onwards (51), so these effects are approximately equivalent to nine years of normal loss of lung function for those with two copies of the coded allele (4 and 7 more than those with one and zero copies respectively). For other SNPs, such as rs2825255, with coded allele (T) frequency of 0.83, association between lung function and air pollutant is observed for certain genotype groups. Using the average NO_2_ measure, those with one and two copies of the effect allele could be subject to reductions in FEV_1_ of approximately 35ml and 75ml (approximately equivalent to 0.75 and 1.5 years of normal lung function decline respectively), as opposed to those with zero copies, where there was no observed statistically significant effect of air pollutant on FEV_1_ (confidence interval overlaps 0).

There were approximately 40,000 individuals with clean lung function data with missing data for education and income status. We expect that those with higher SES and higher income are more likely to have complete data thus the data is not missing at random. We did not carry out imputation as it is difficult to know which might introduce more bias, imputation or exclusion and thus carried out a complete-case analysis. Further studies are required in this respect. Previous studies have reported modification of air pollution effects on lung function when considering SES (4, 52–54) possibly due to differences in housing conditions, indoor air quality, nutrition and occupation (54). Adjusting for SES and presenting interaction effects across educational and income groups did not produce a notable modification of interaction effects in our analyses, suggesting that observed differences in the effect of air pollution across genotype groups are not mediated or confounded by socio-economic status.

There are other limitations with this study. We only had air pollution data at baseline with some limitations in the availability and did not have follow-up data. An analysis of a German cohort of 601 elderly women (mainly non-smokers) with three follow-ups from 1985-2013 suggested that changes in air pollution over time was associated with improvements in lung function, modified by genetic factors (55). In addition, there are limitations with the ESCAPE models (8, 9). Exposure estimates are based on place of residence so will not capture variability in exposure related to work and leisure activities outside the home, which may have led to exposure misclassification bias making it harder to detect effects. Furthermore, it must be noted that our analysis includes imputed genetic dosages alongside directly genotyped data and we only considered an additive genetic model for our analysis. Previous studies for certain antioxidant gene SNPs such as rs1695 in *GSTP1* have also considered the suitability of alternative genetic models (56, 57).

## Conclusions

We have identified genetic variants whose effect on lung function is dependent on air pollution exposure levels. This could help identify high-risk genetic subgroups whose lung function could be more susceptible to the effects of outdoor air pollution. While this is the largest study of this type to date, we highlight the need for replication in independent datasets with recorded lung function, for which availability is currently limited. We hope that future replication and further biological studies of gene function will help to establish the genes and biological pathways involved.

## Data Availability

The data will be made available following peer review.

## Funding and acknowledgements

Louise Wain holds a GSK/British Lung Foundation Chair in Respiratory Research (C17-1). The research was partially supported by the National Institute for Health Research (NIHR) Leicester Biomedical Research Centre; the views expressed are those of the author(s) and not necessarily those of the National Health Service (NHS), the NIHR or the Department of Health.

Martin Tobin is supported by a Wellcome Trust Investigator Award (WT202849/Z/16/Z) and holds an NIHR Senior Investigator Award.

Anna Hansell acknowledges funding from the Health Protection Research Unit in Environmental Exposures and Health, a partnership between Public Health England, the Health and Safety Executive and the University of Leicester. The views expressed are those of the authors and not necessarily those of the NHS the NIHR, Public Health England, the Health and Safety Executive or the Department of Health and Social Care.

The analysis was undertaken using UK Biobank data application 648.

This research used the High Performance Computing facilities at the University of Leicester (ALICE and SPECTRE).

