## Supplementary_Figures_1-9 for "Genome-wide gene-air pollution interaction analysis of lung function in 300,000 individuals"

Supplementary figure 1: SNP QC flow chart

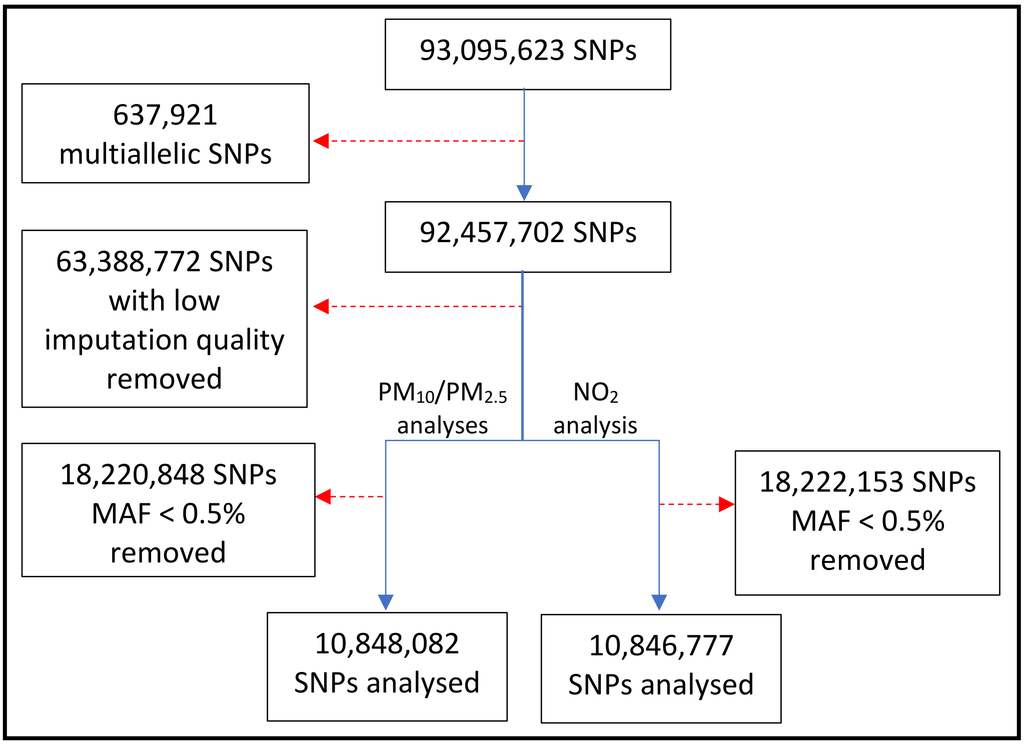

Supplementary figure 2: QQ plots for GWAS (with genomic inflation factor, λ)

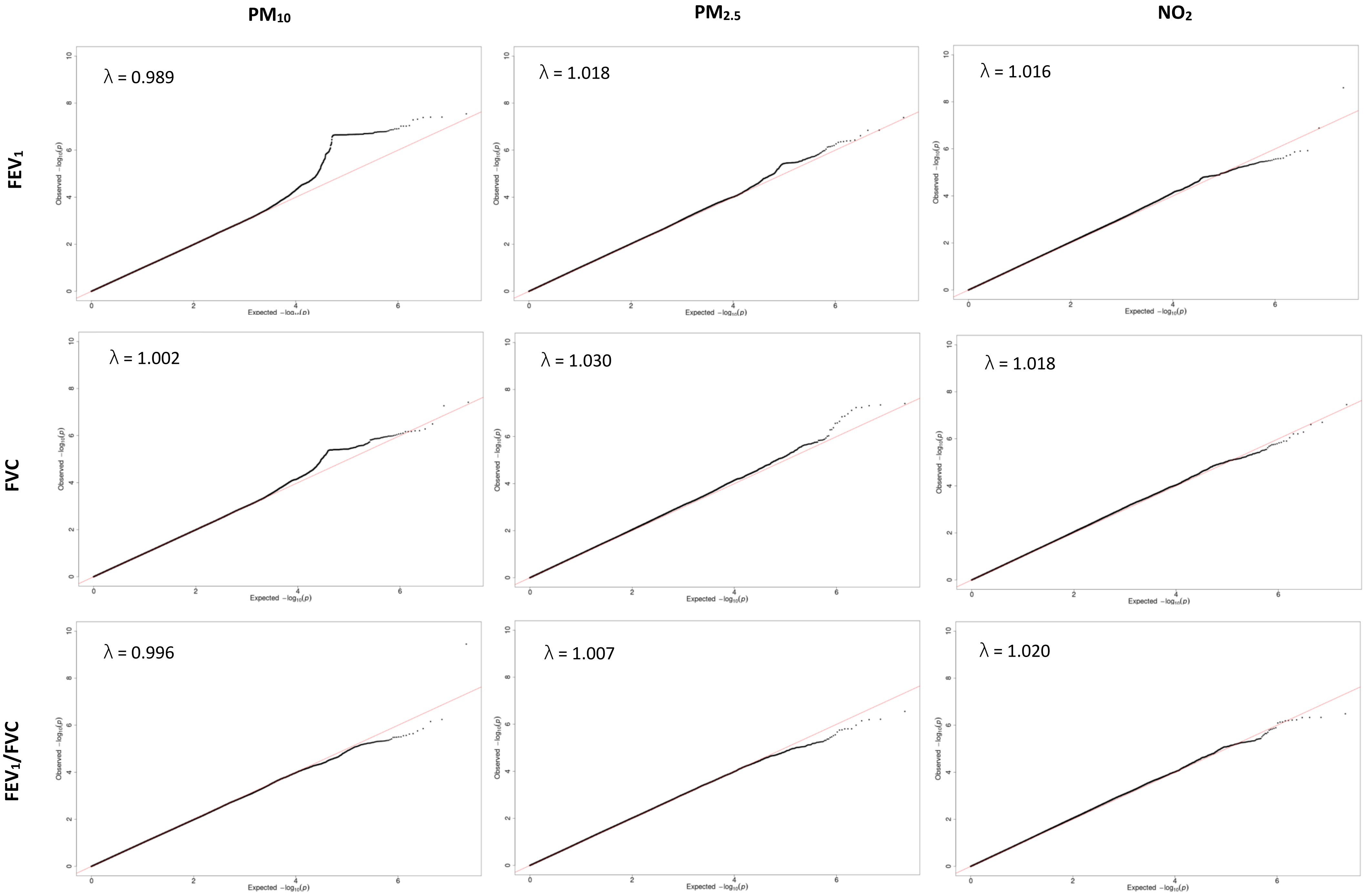
PM10
PM2.5
NO2
FEV1
FVC
FEV1/FVC
λ = 0.989
λ = 1.018
λ = 1.016
λ = 1.002
λ = 1.030
λ = 1.018
λ = 0.996
λ = 1.007
λ = 1.020
Observed −log10(p)
Expected −log10(p)

#### Supplementary figure 3: Region plots for genome-wide signals

### 1) rs74048016

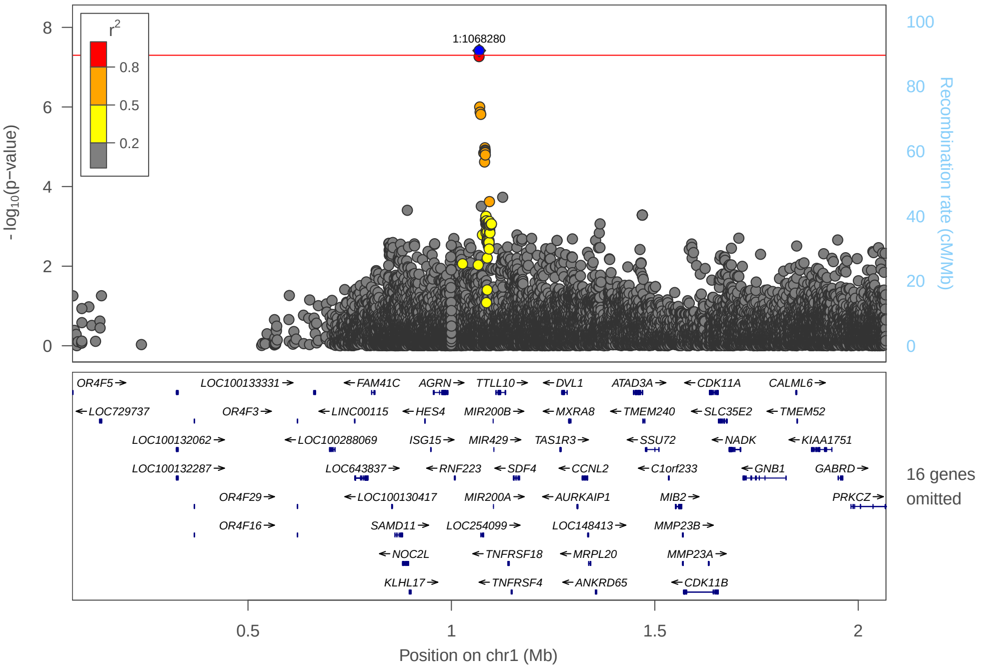

### 2) rs28666788

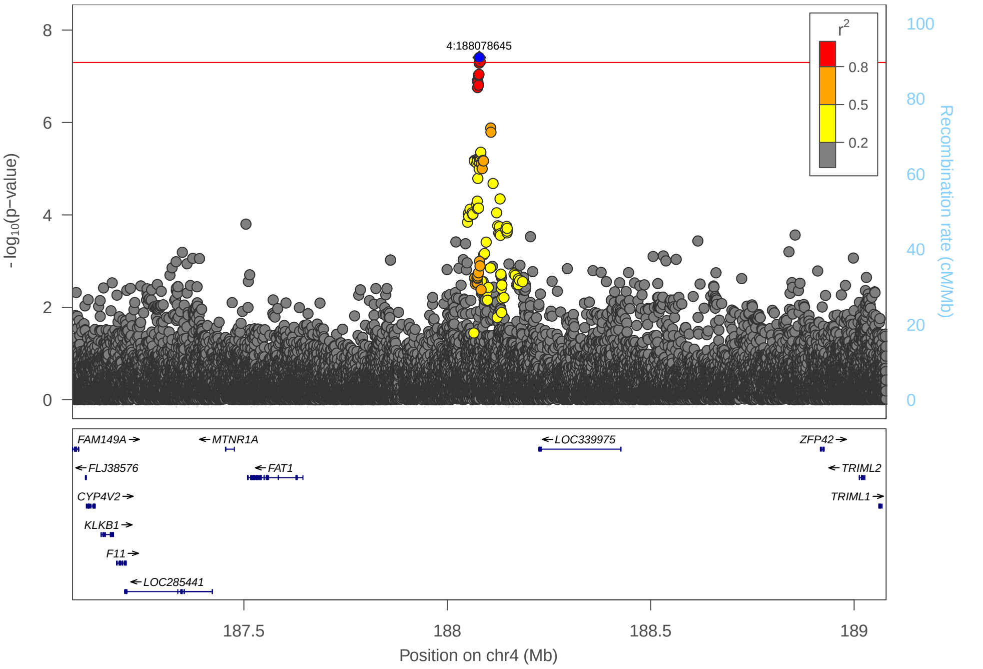

3) rs192415220

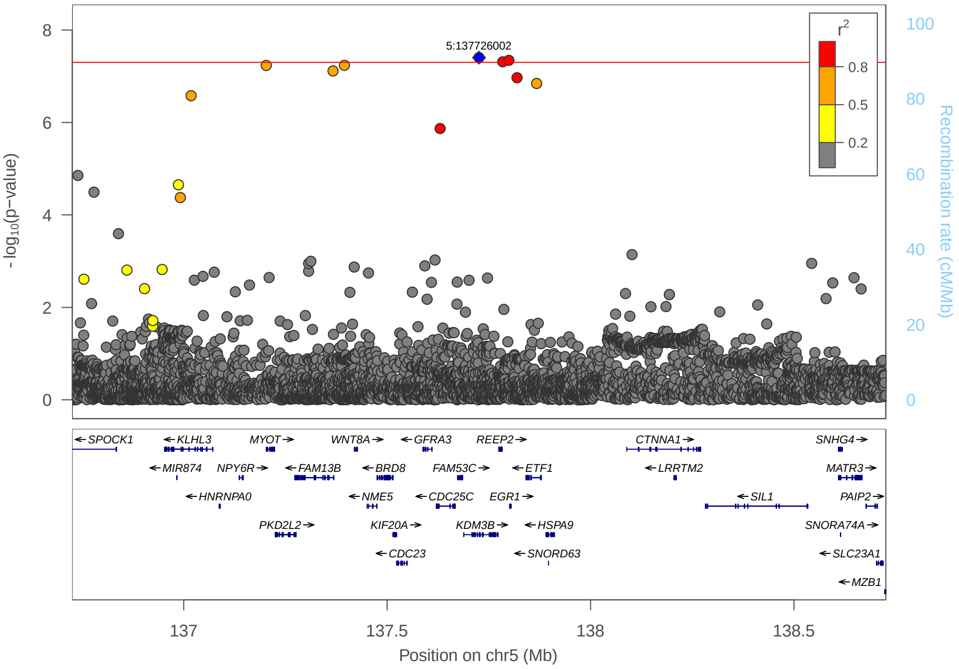

4) rs137914543

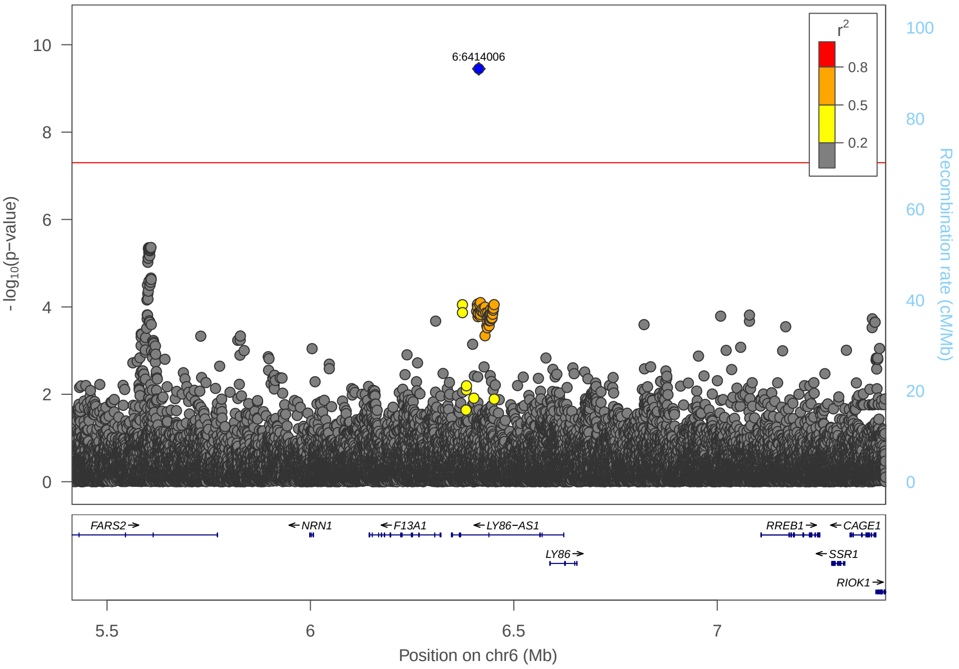

5) rs138235384

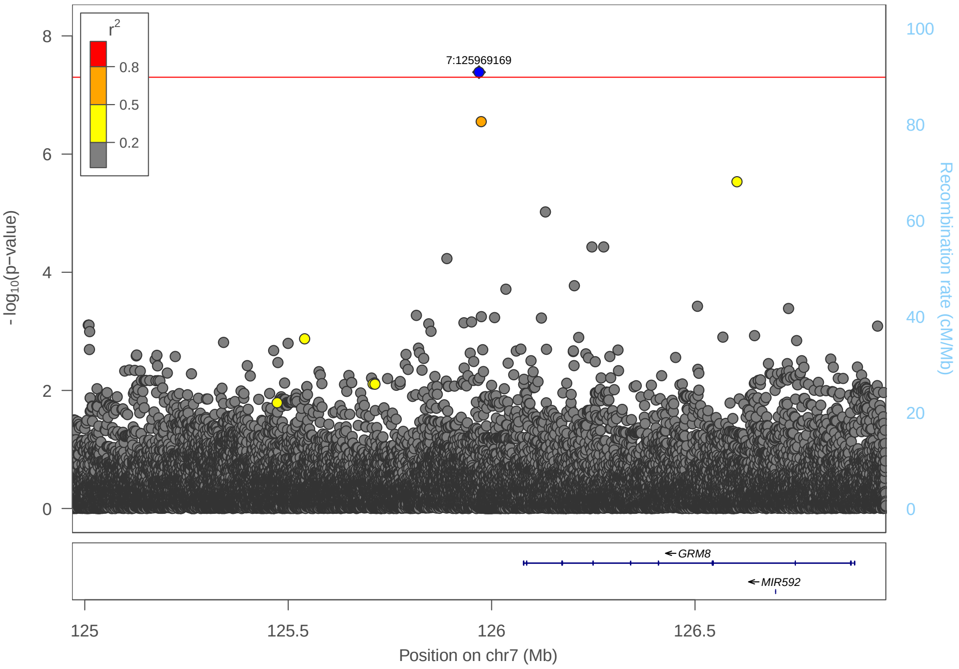

6) rs762101031

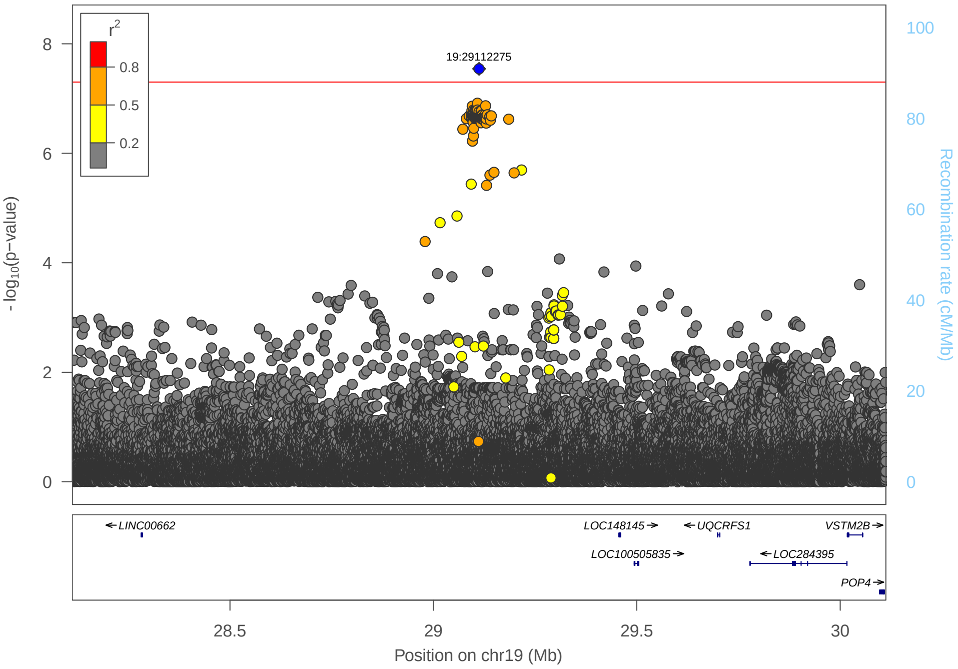

7) rs2825255 ($FEV_1$)

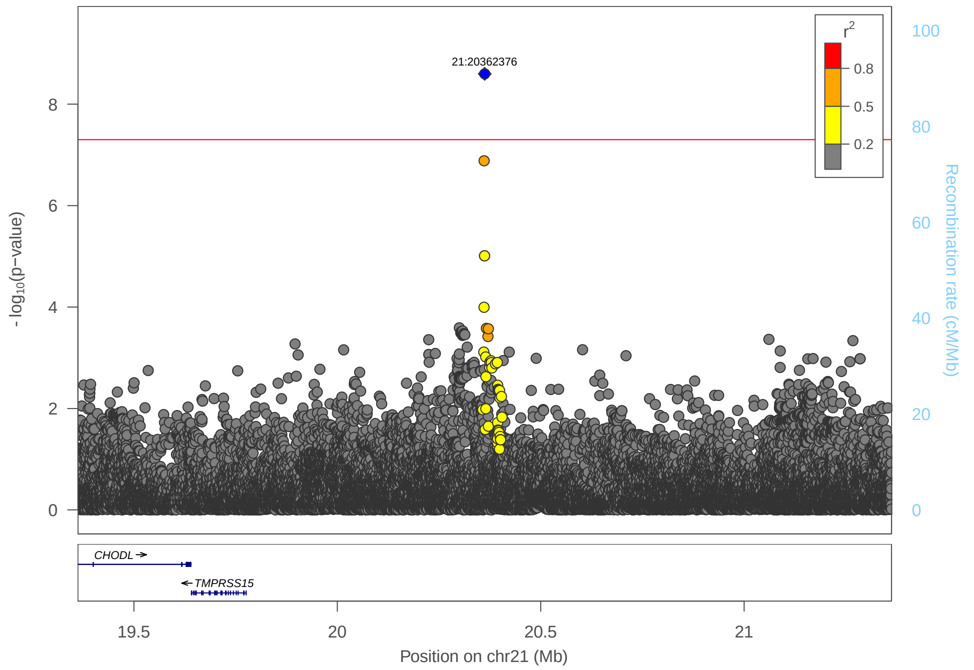

8) rs2825255 (FVC)

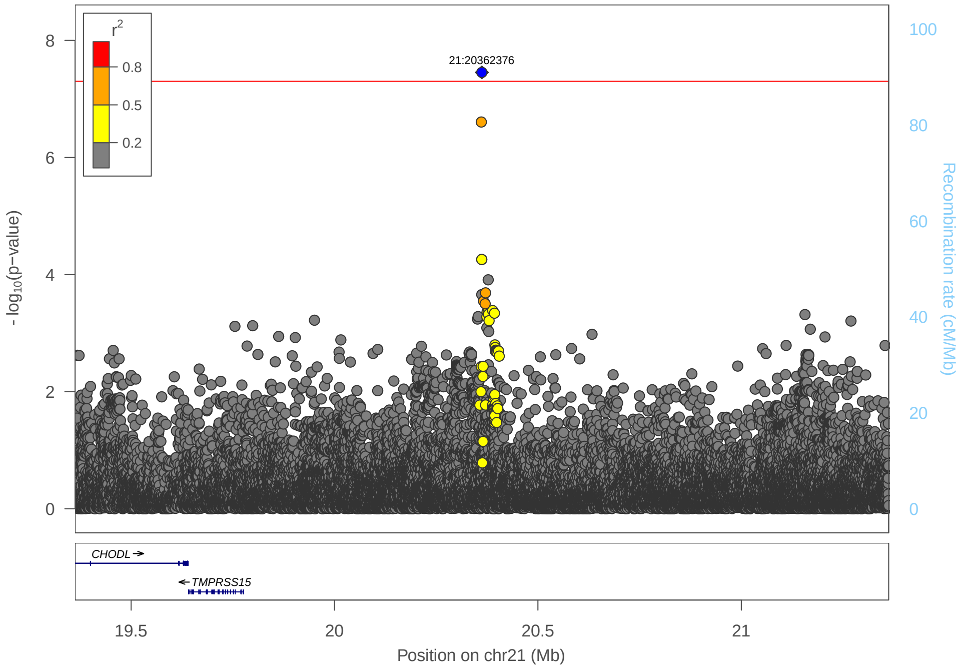

Supplementary figure 4: Interaction plots for genome-wide signals

1) rs74048016

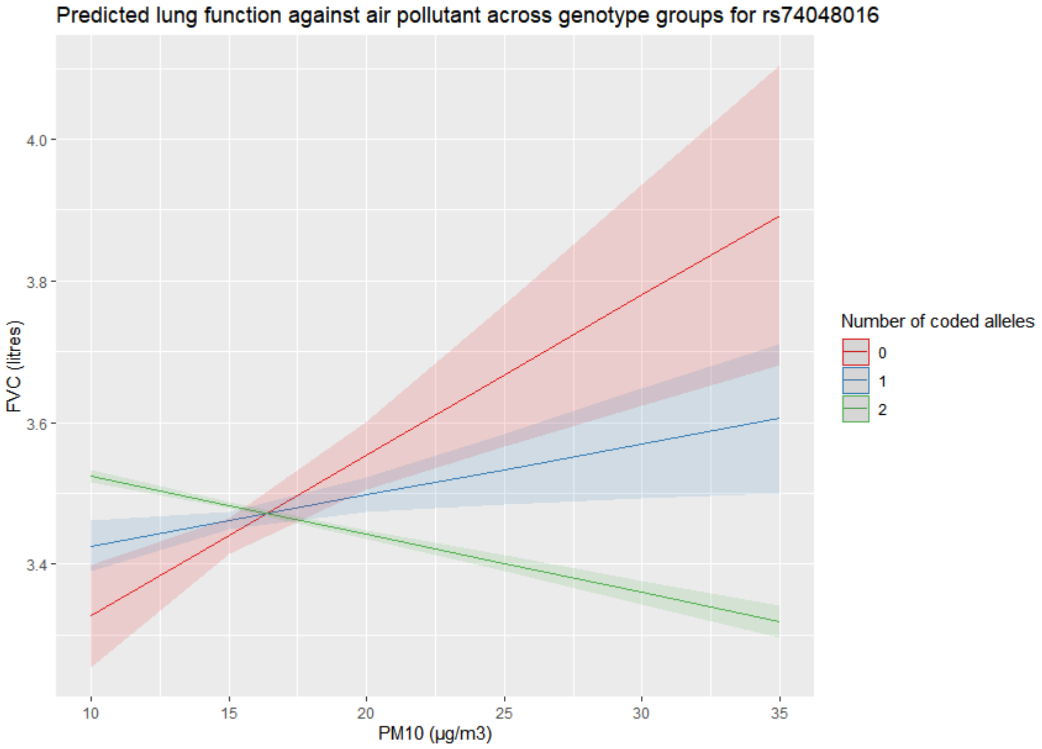

2) rs28666788

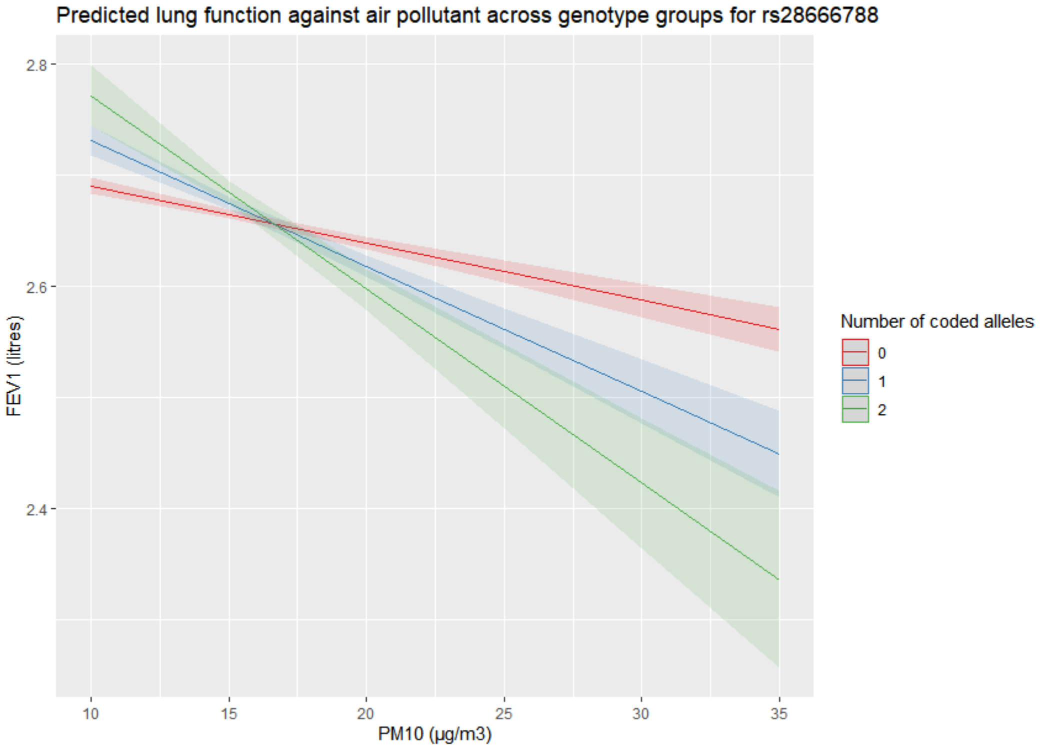

3) rs192415220

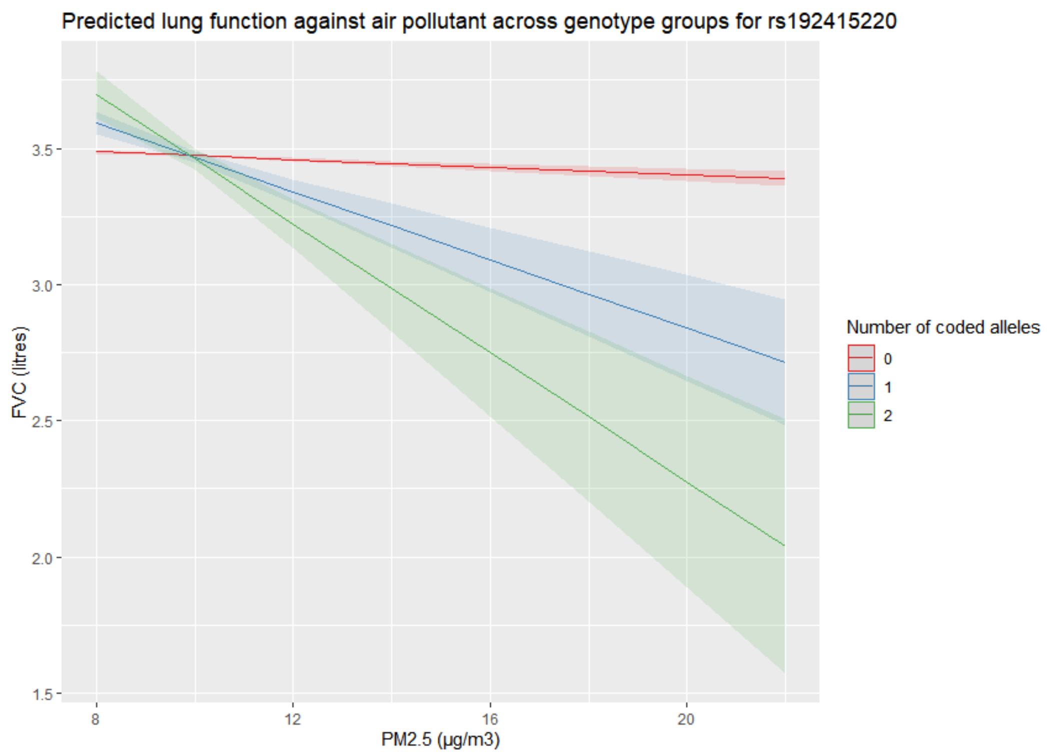

4) rs137914543

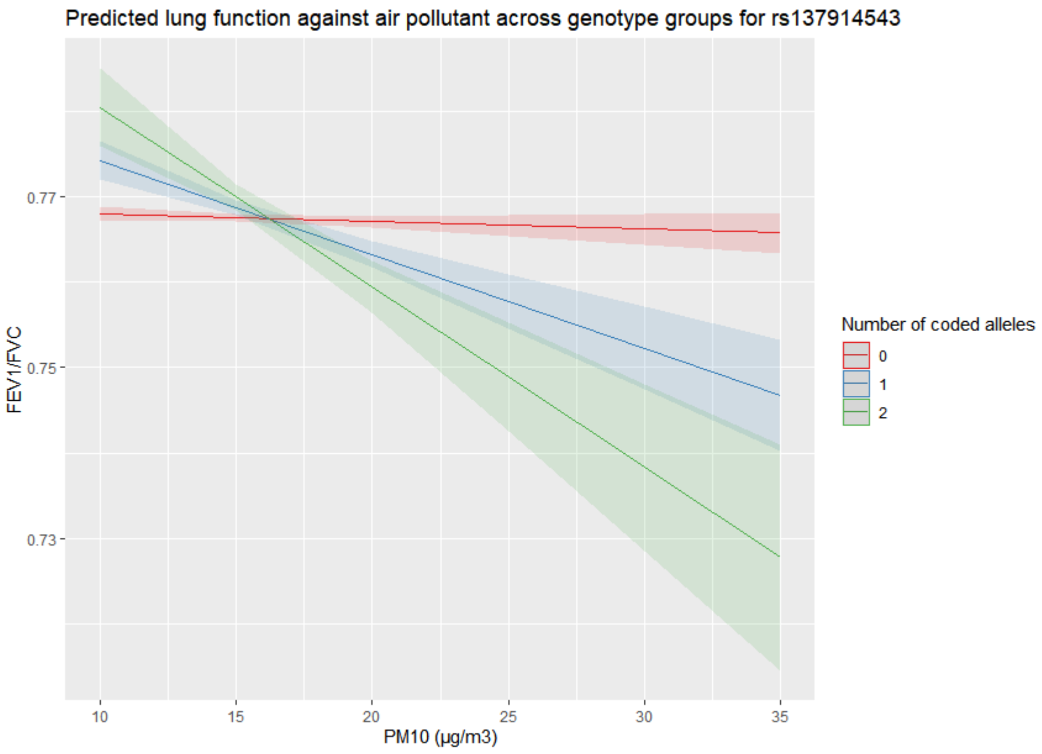

5) rs138235384

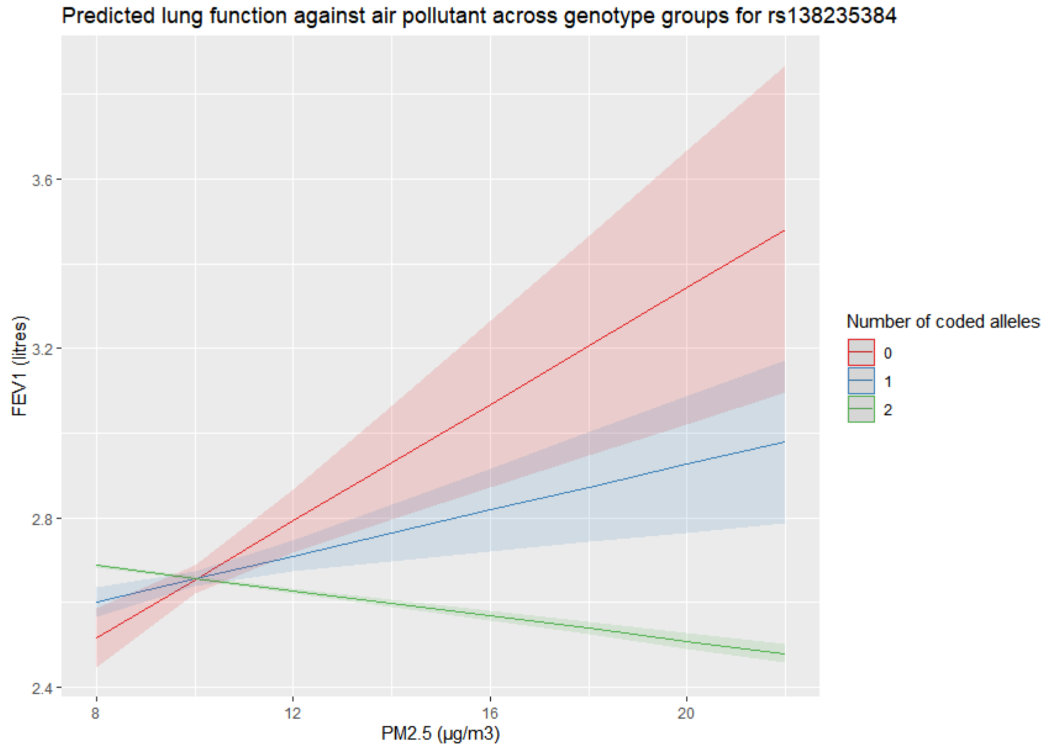

6) rs762101031

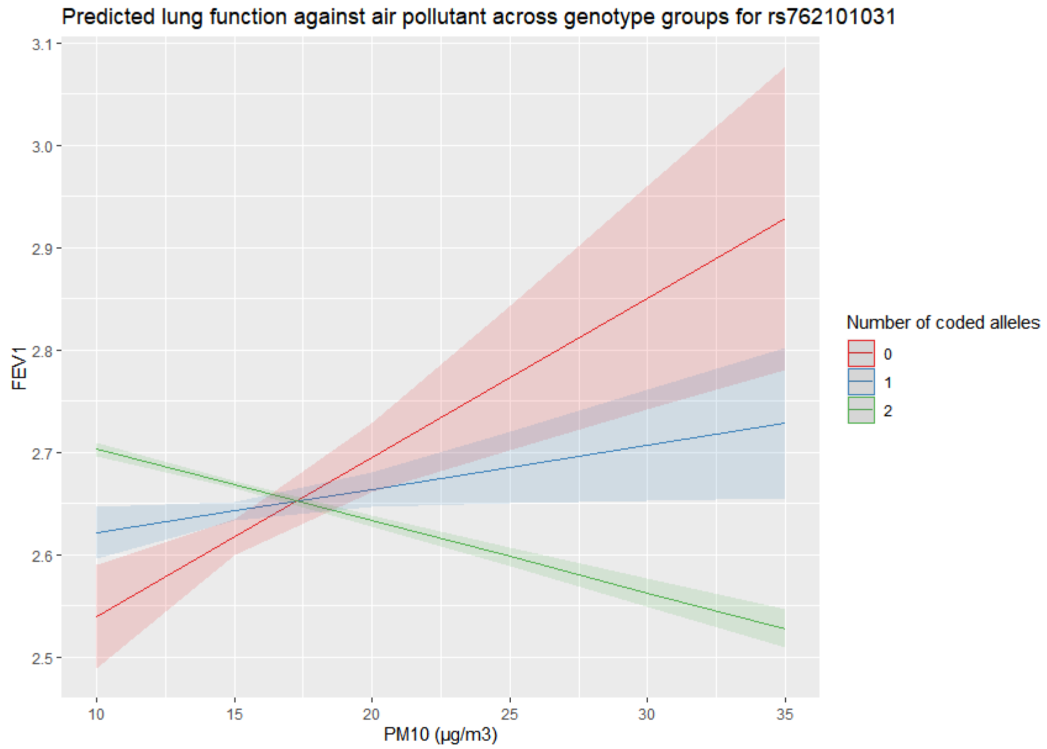

7) rs2825255 ($FEV_1$)

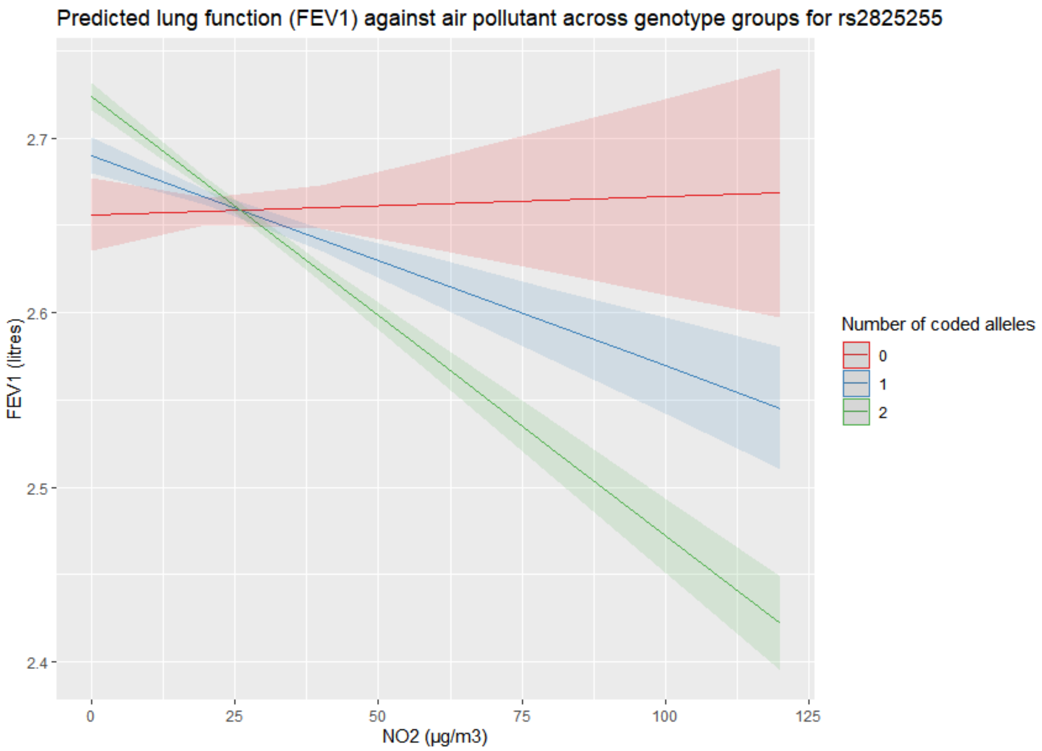

8) rs2825255 (FVC)

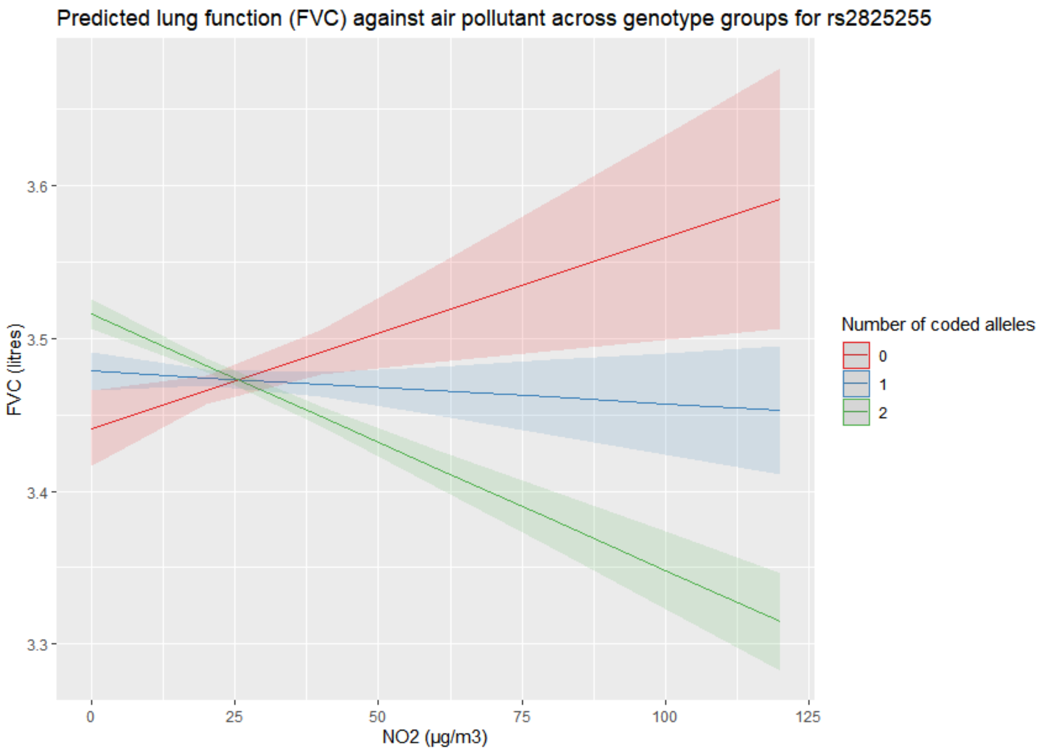

### Supplementary figure 5: Region plots for suggestive signals

## 1) rs140250292

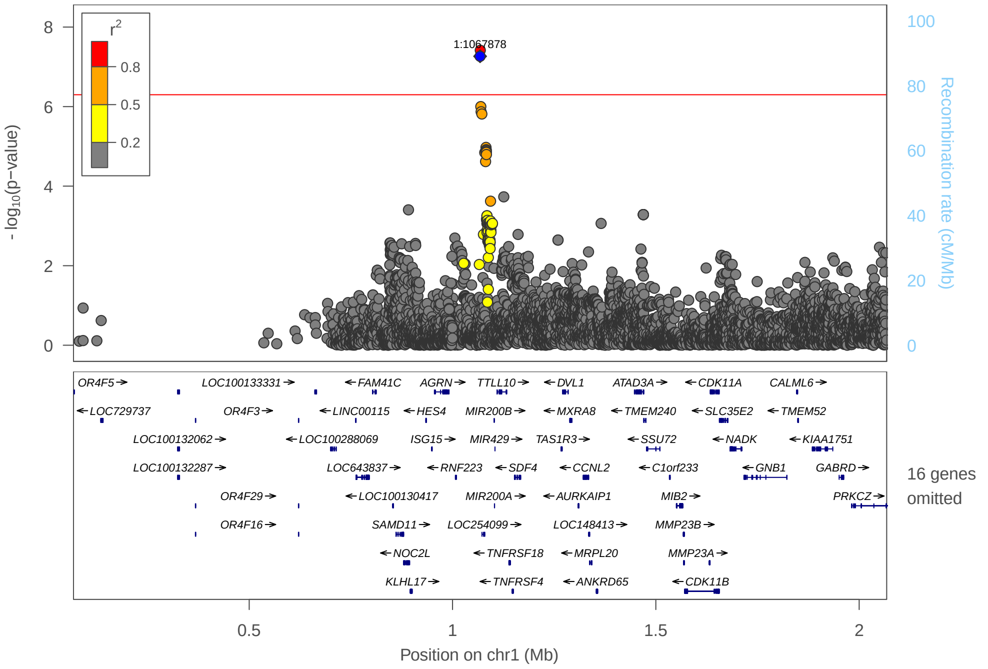

## 2) rs35380252

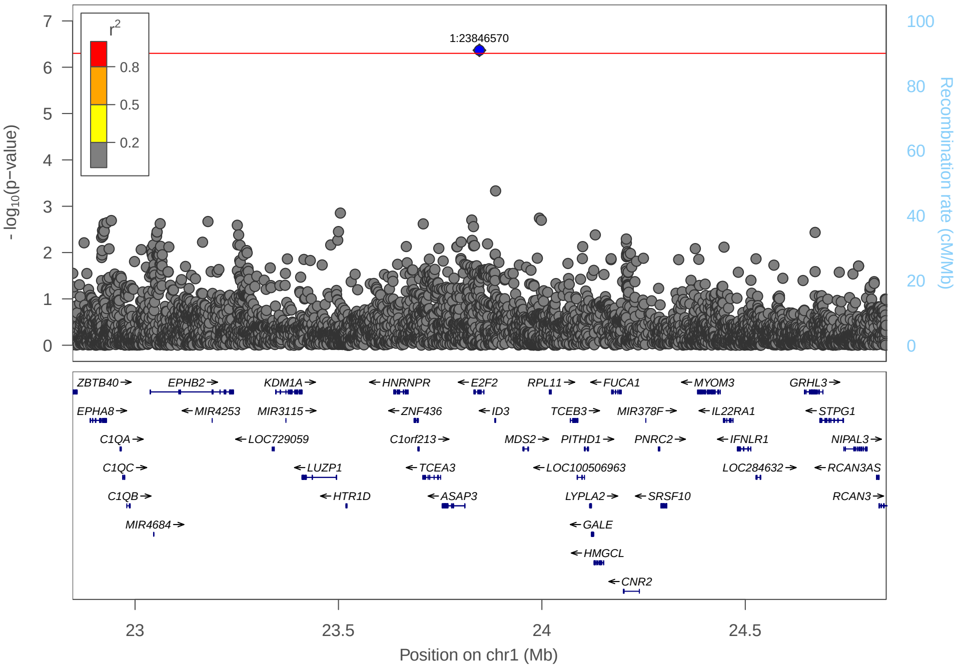

3) rs6661026

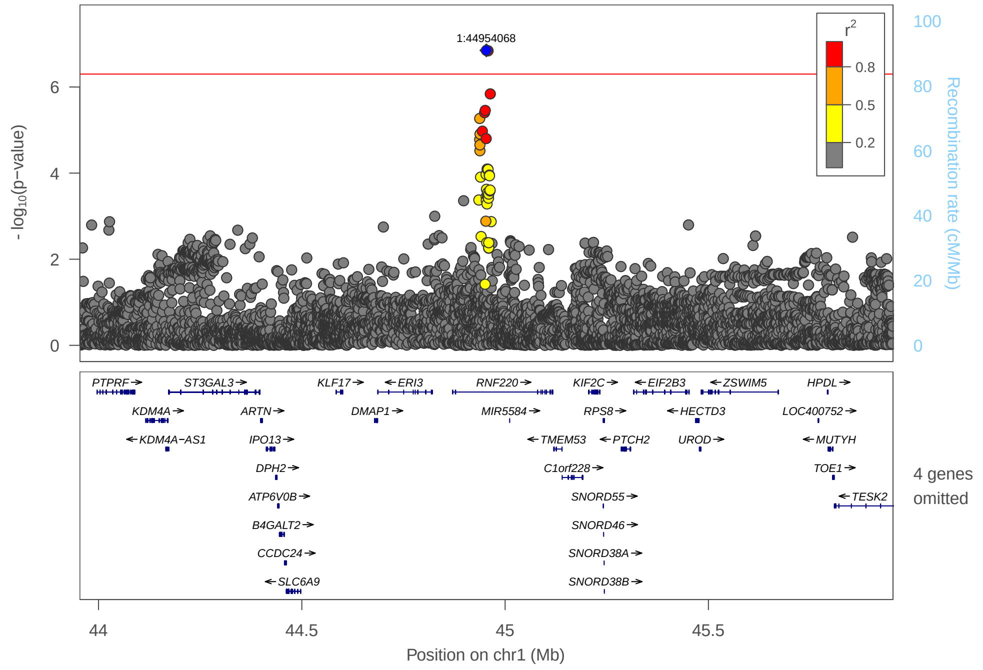

4) rs10082259

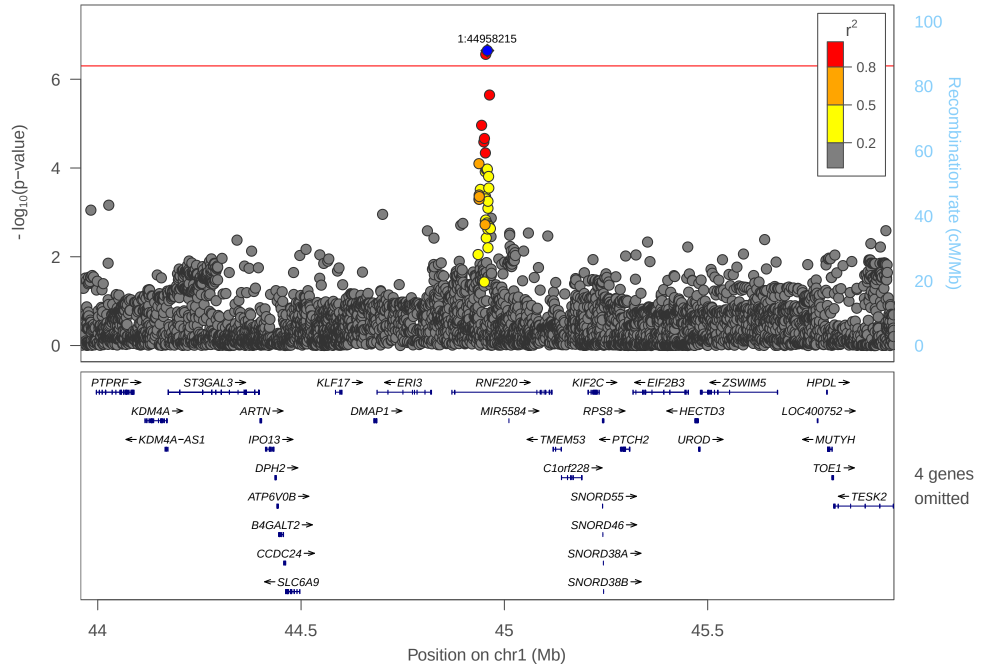

5) rs769937512 (FEV$_1$, PM$_{10}$)

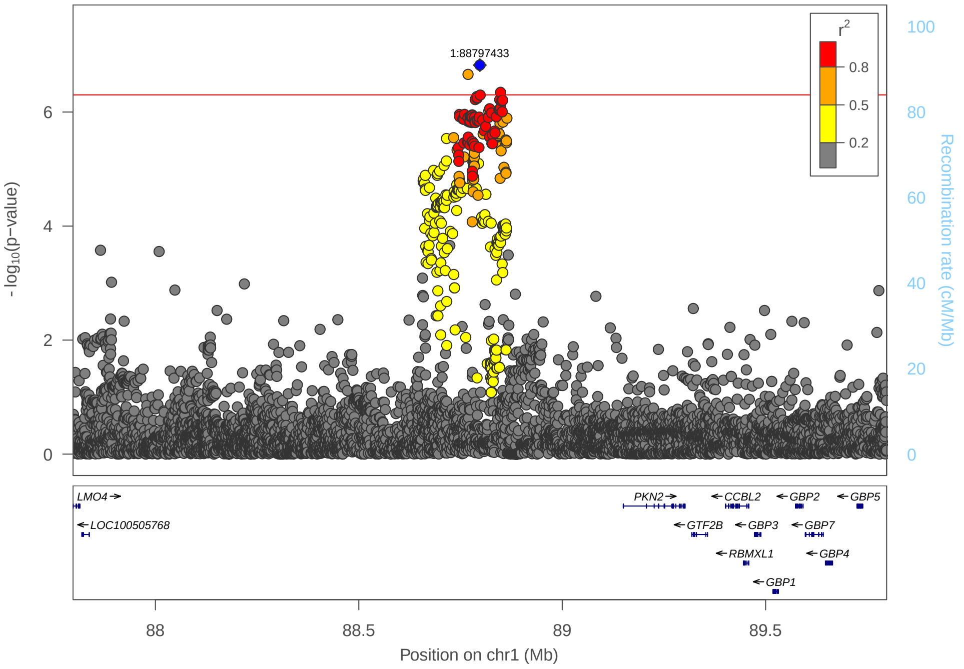

6) rs769937512 (FVC, PM$_{10}$)

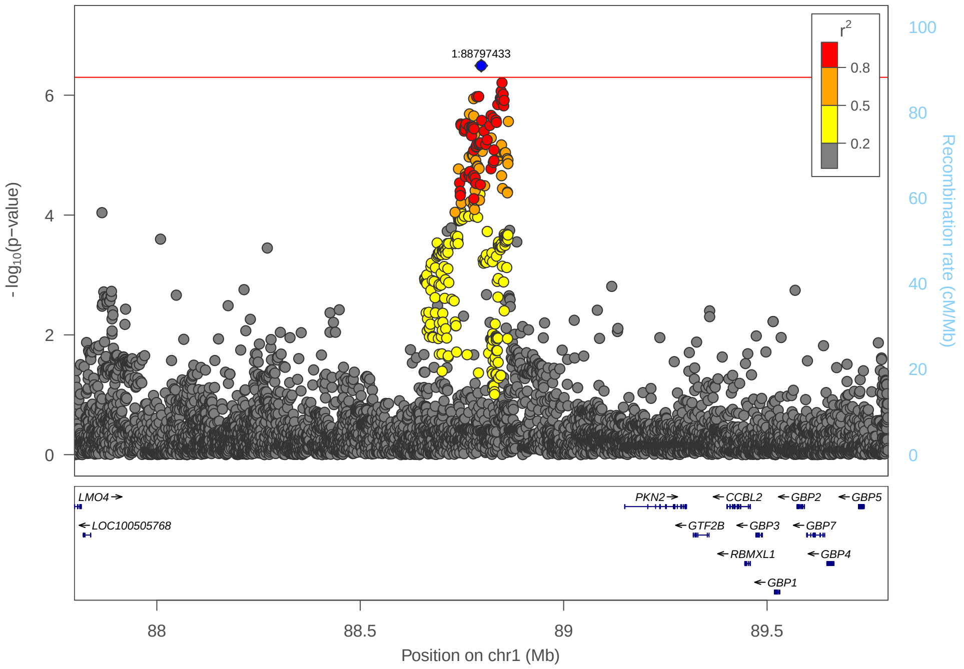

7) rs11677115

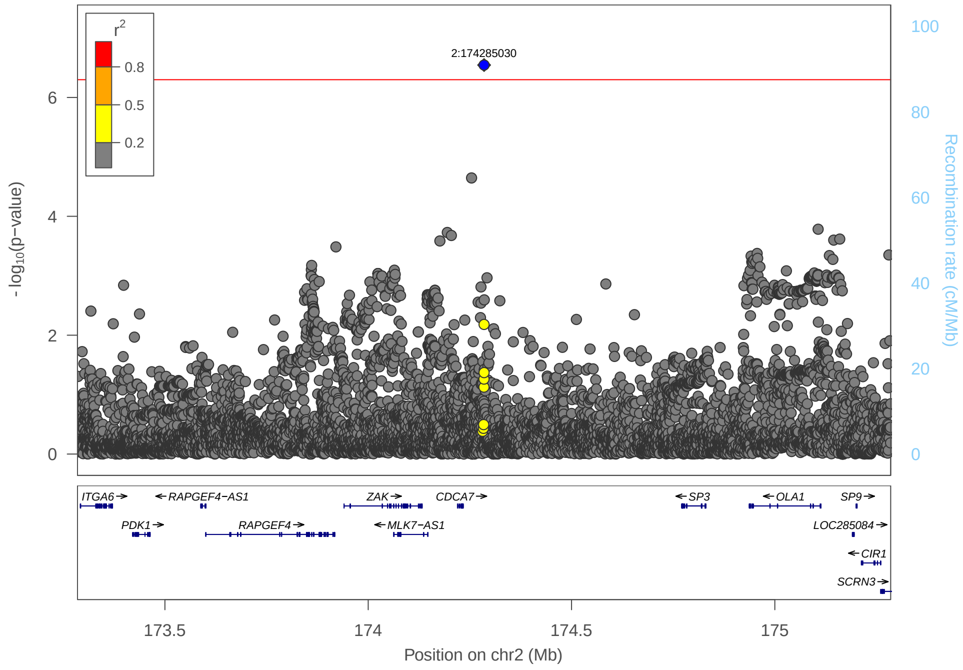

8) 3:322596_CCACA_C

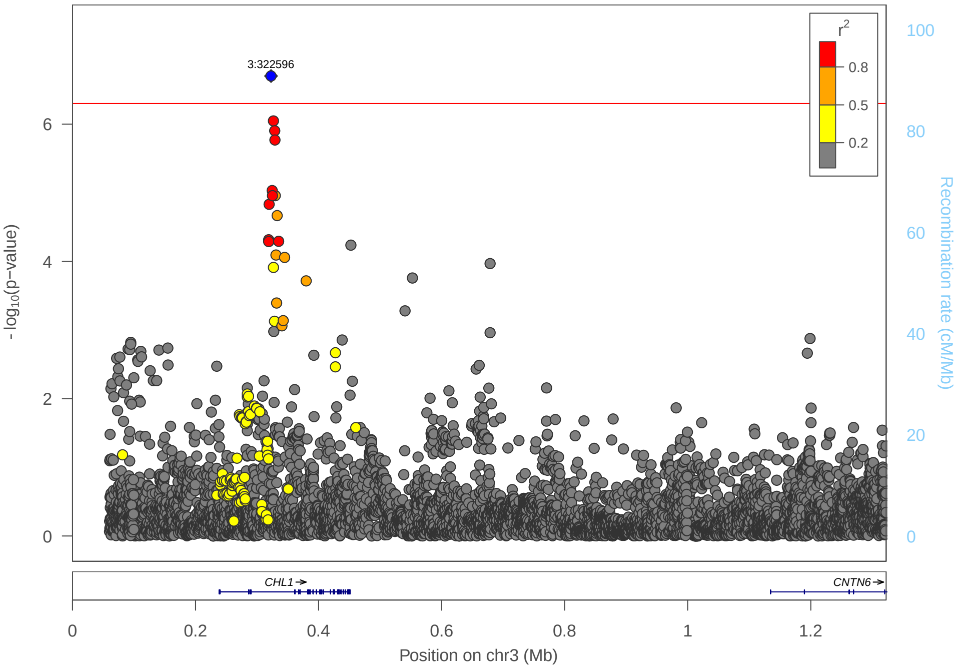

9) rs200460259

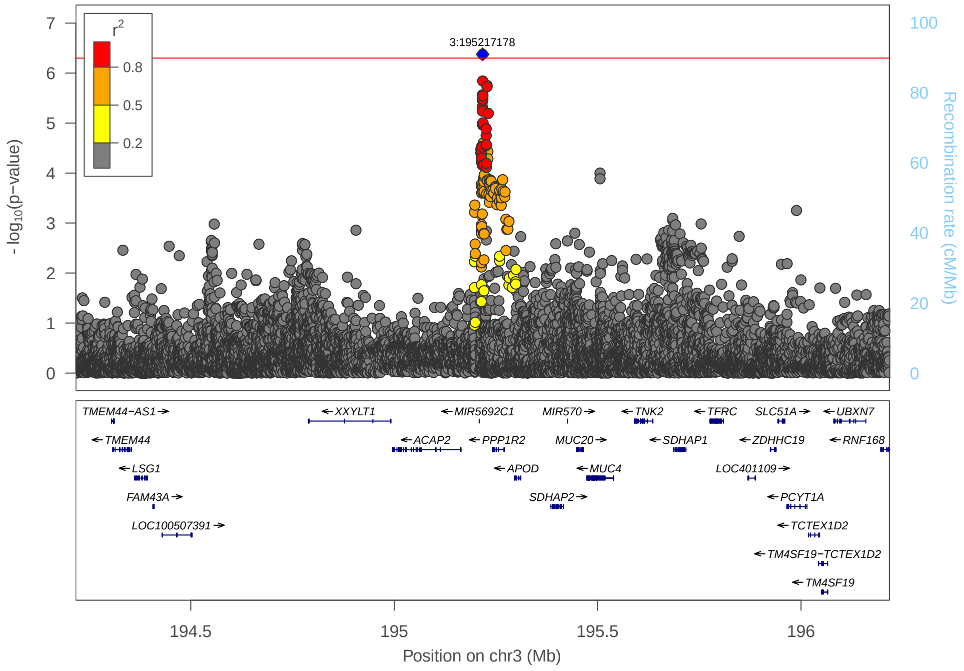

10) rs111552599

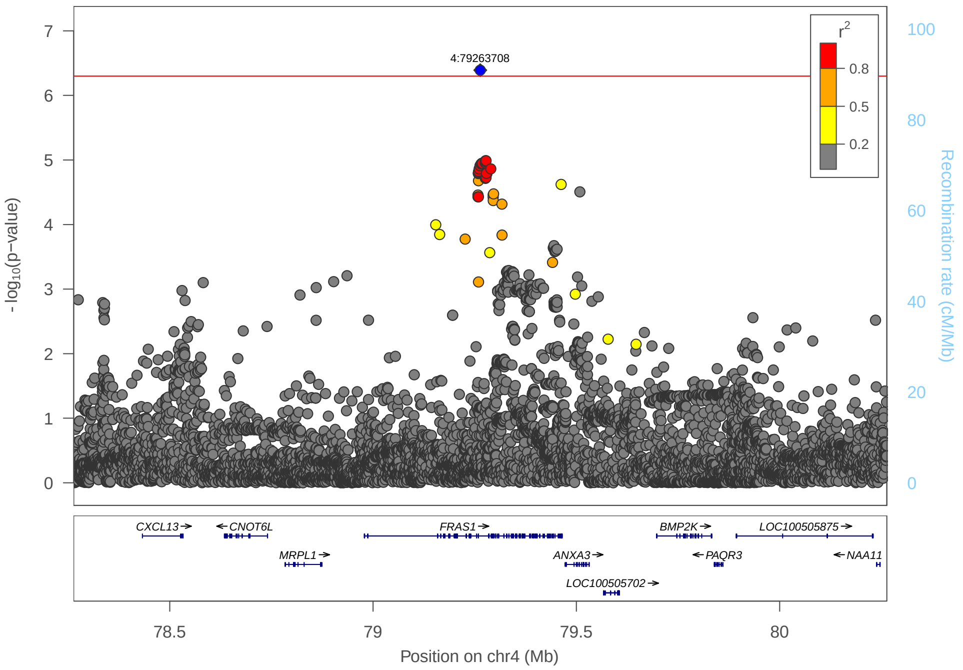

## 11) rs28665554

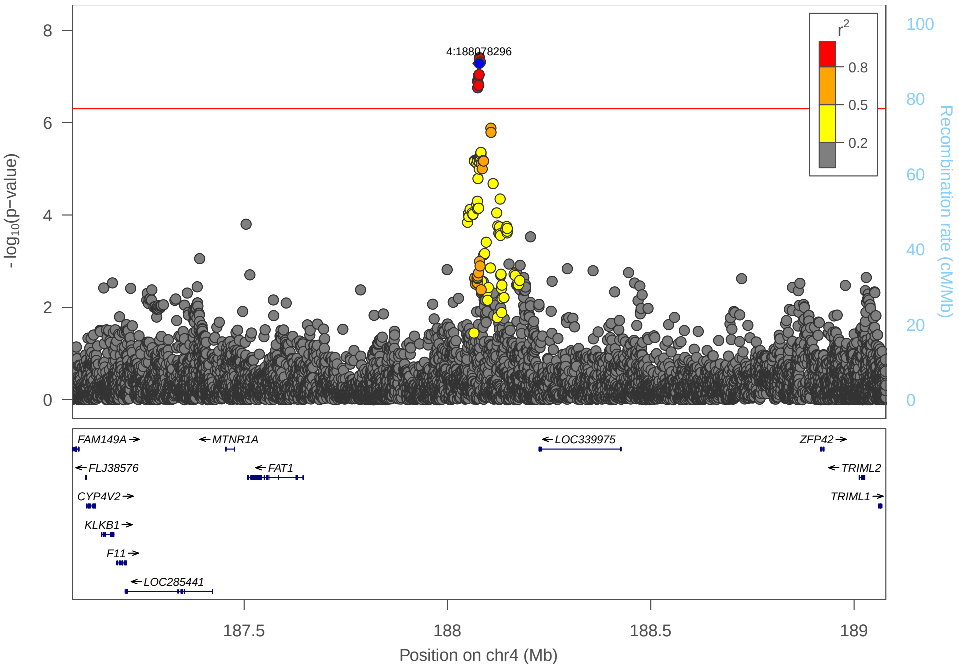

## 12) rs189103140

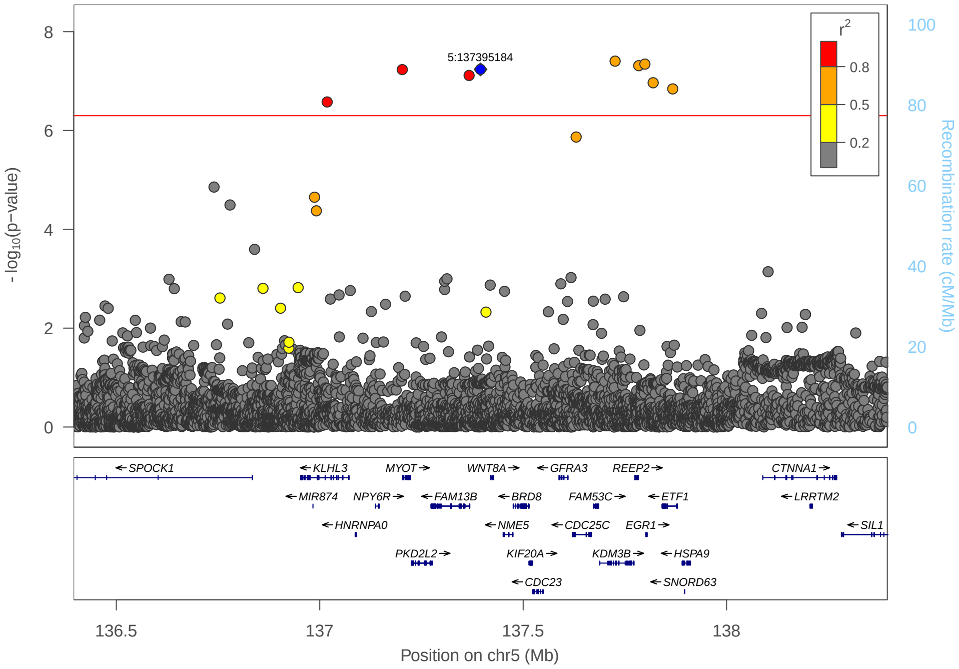

## 13) rs192415220

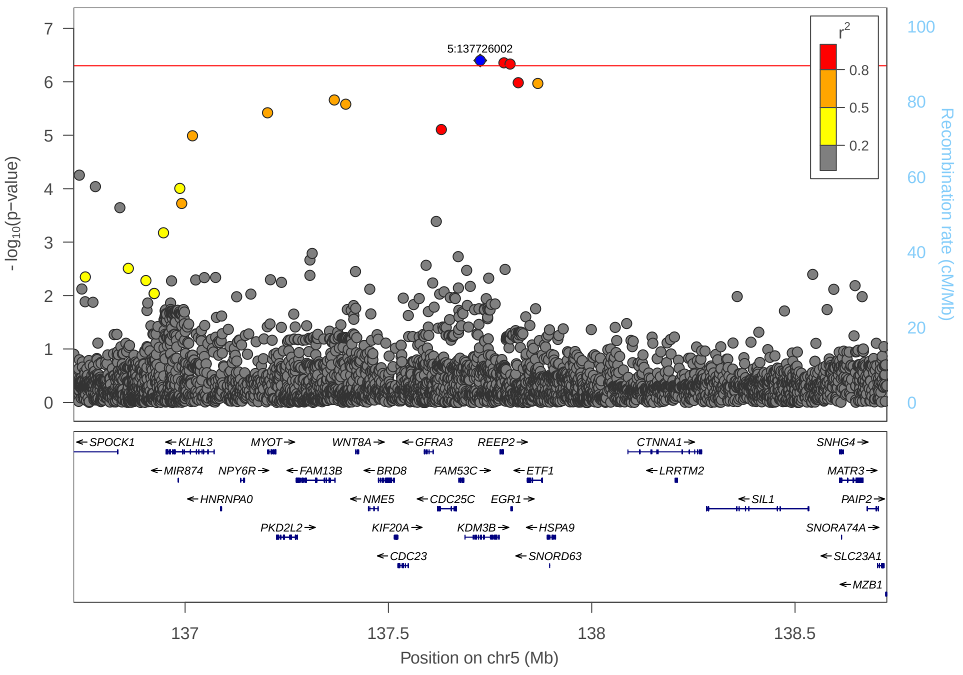

## 14) rs138235384

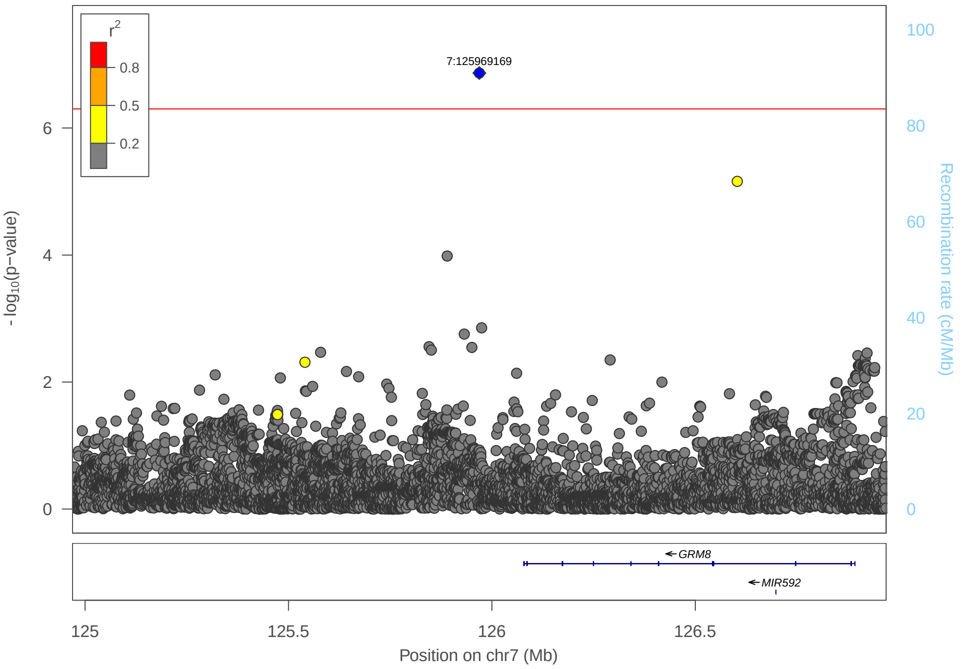

15) rs73163133

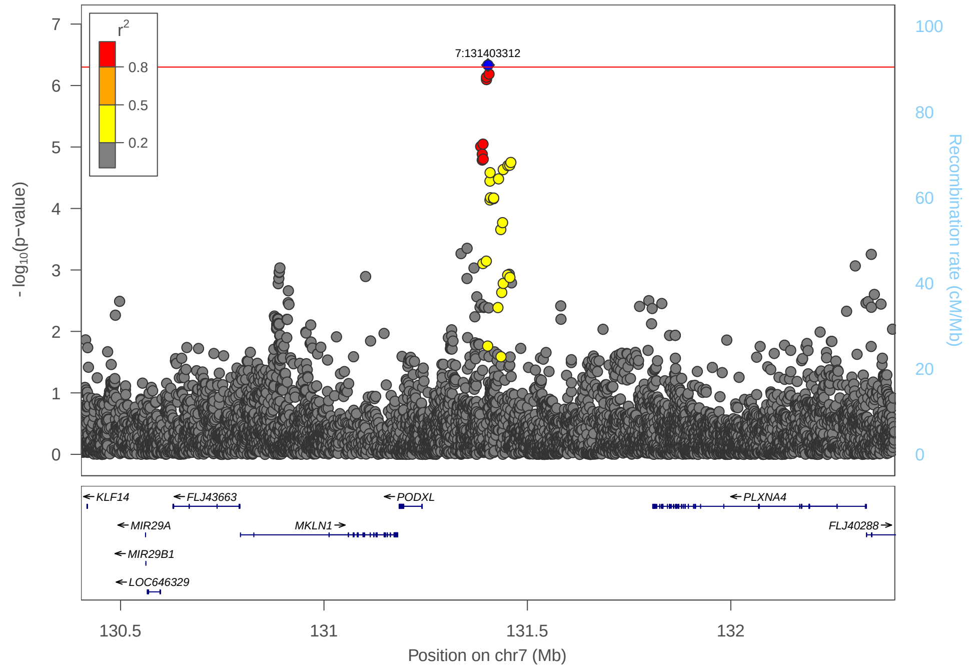

16) rs139556451

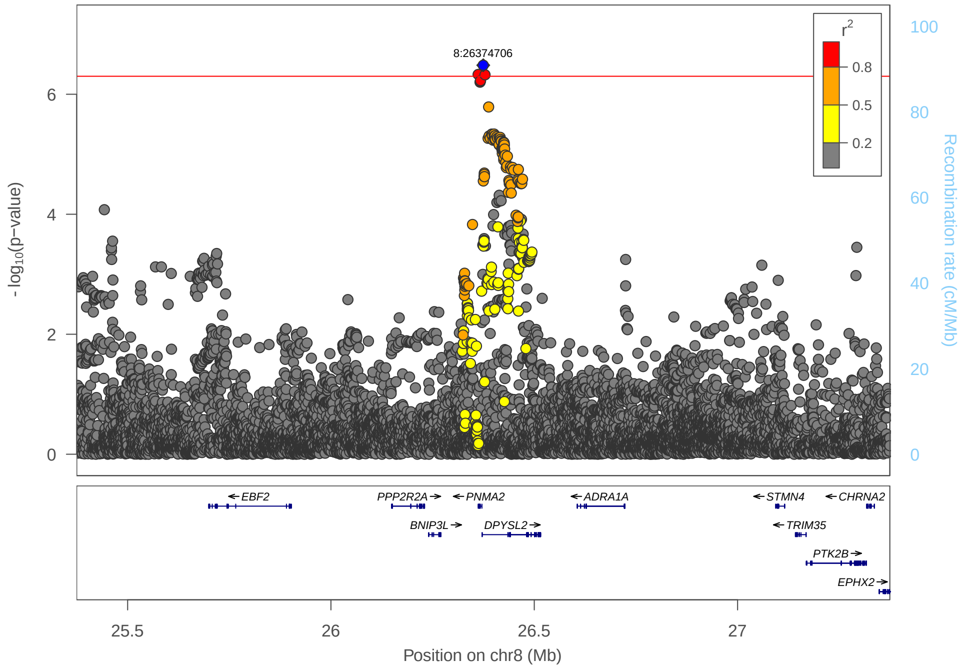

17) rs7098338

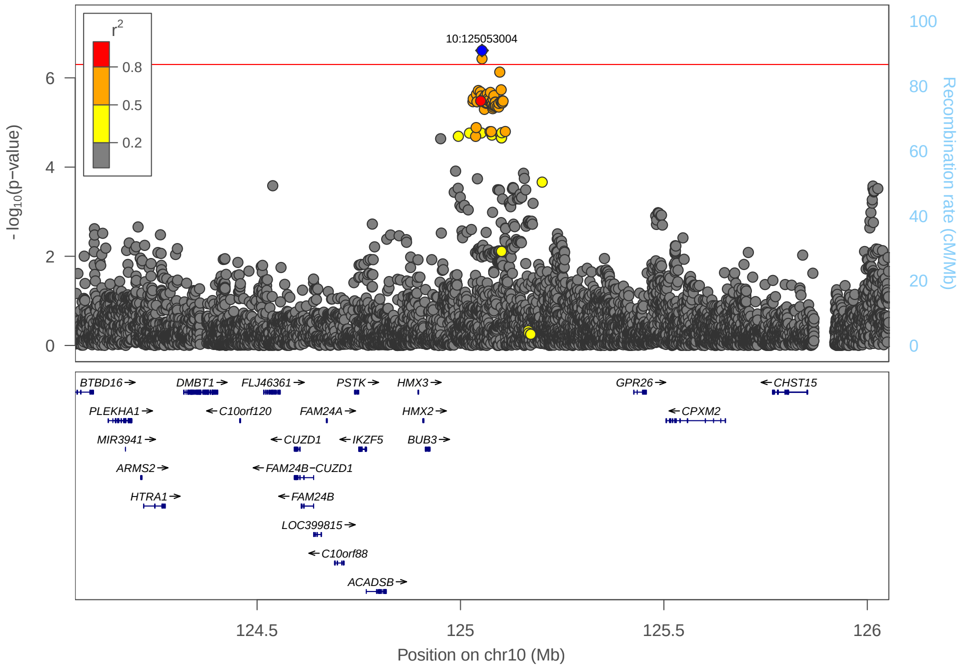

18) 19:29108143_GA_G

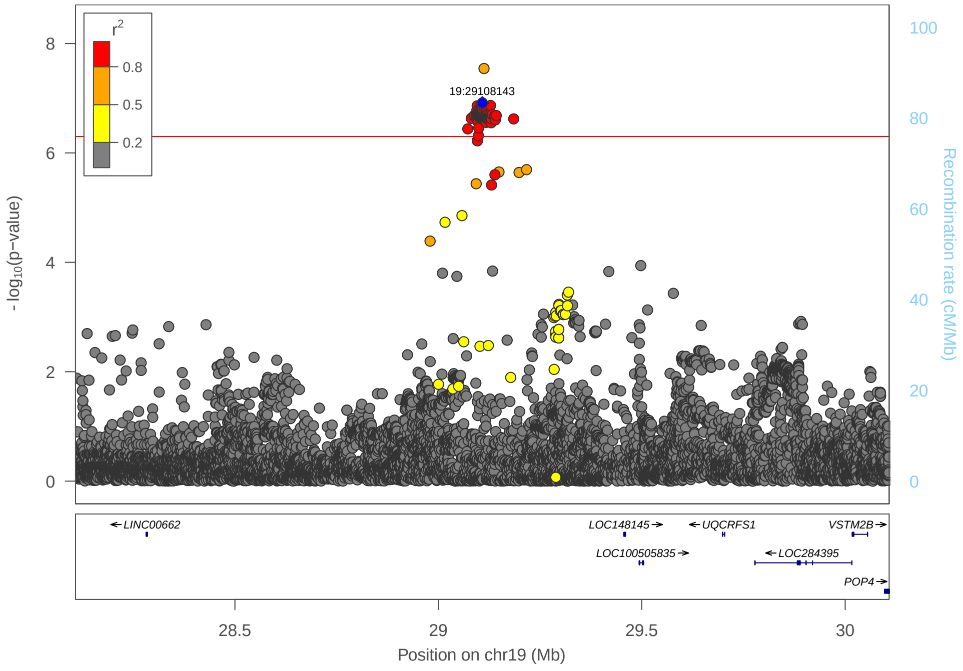

#### 19) rs111676952 ($FEV_1$, $NO_2$)

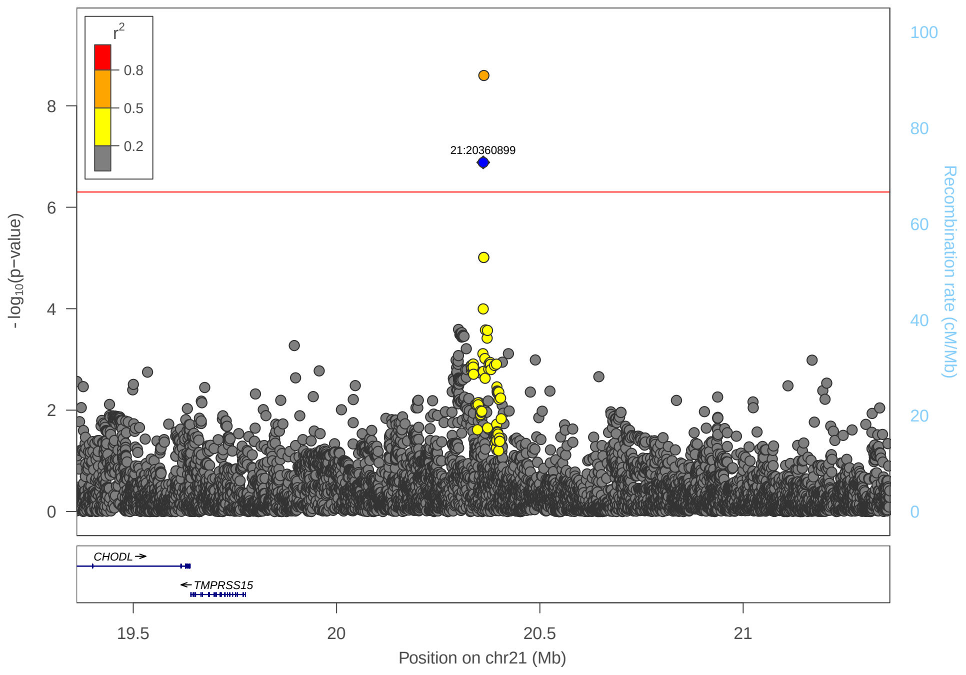

#### 20) rs111676952 (FVC, $NO_2$)

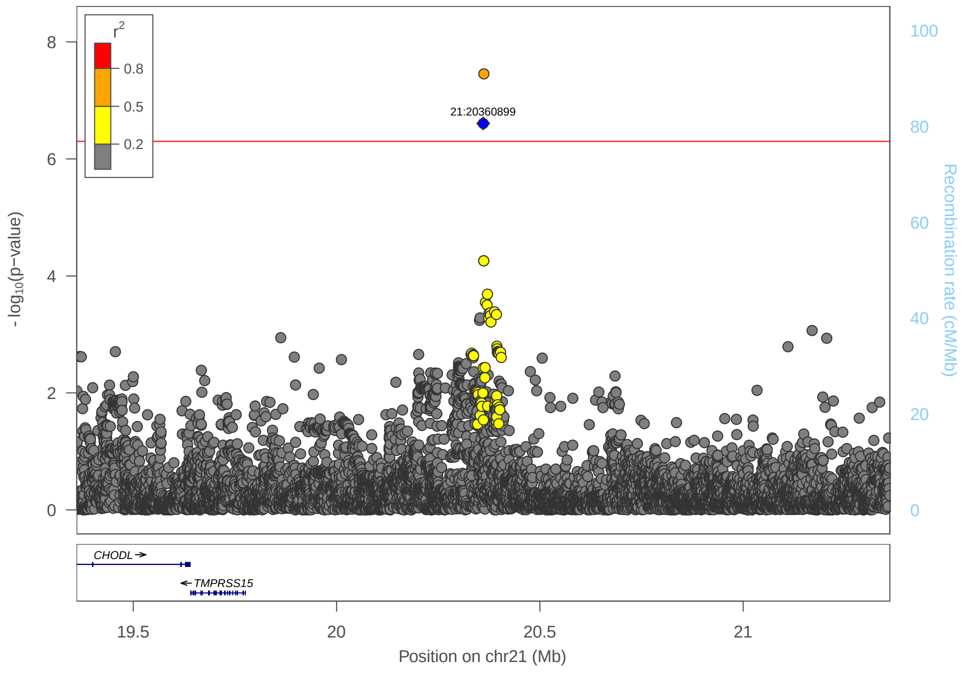

Supplementary figure 6: GARFIELD analysis (FVC, $NO_2$)

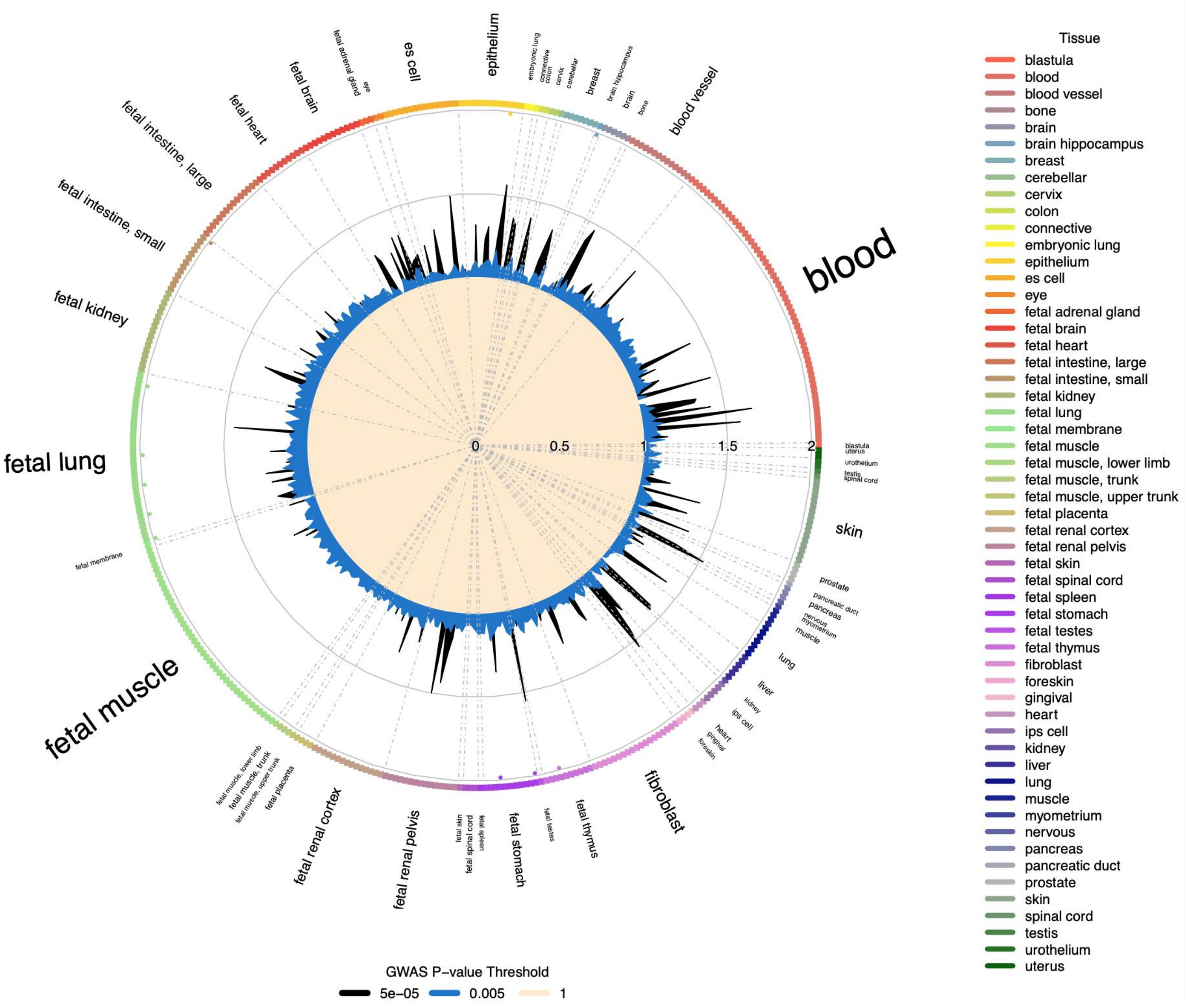

Supplementary figure 7: Effect of SES adjustment on top signals

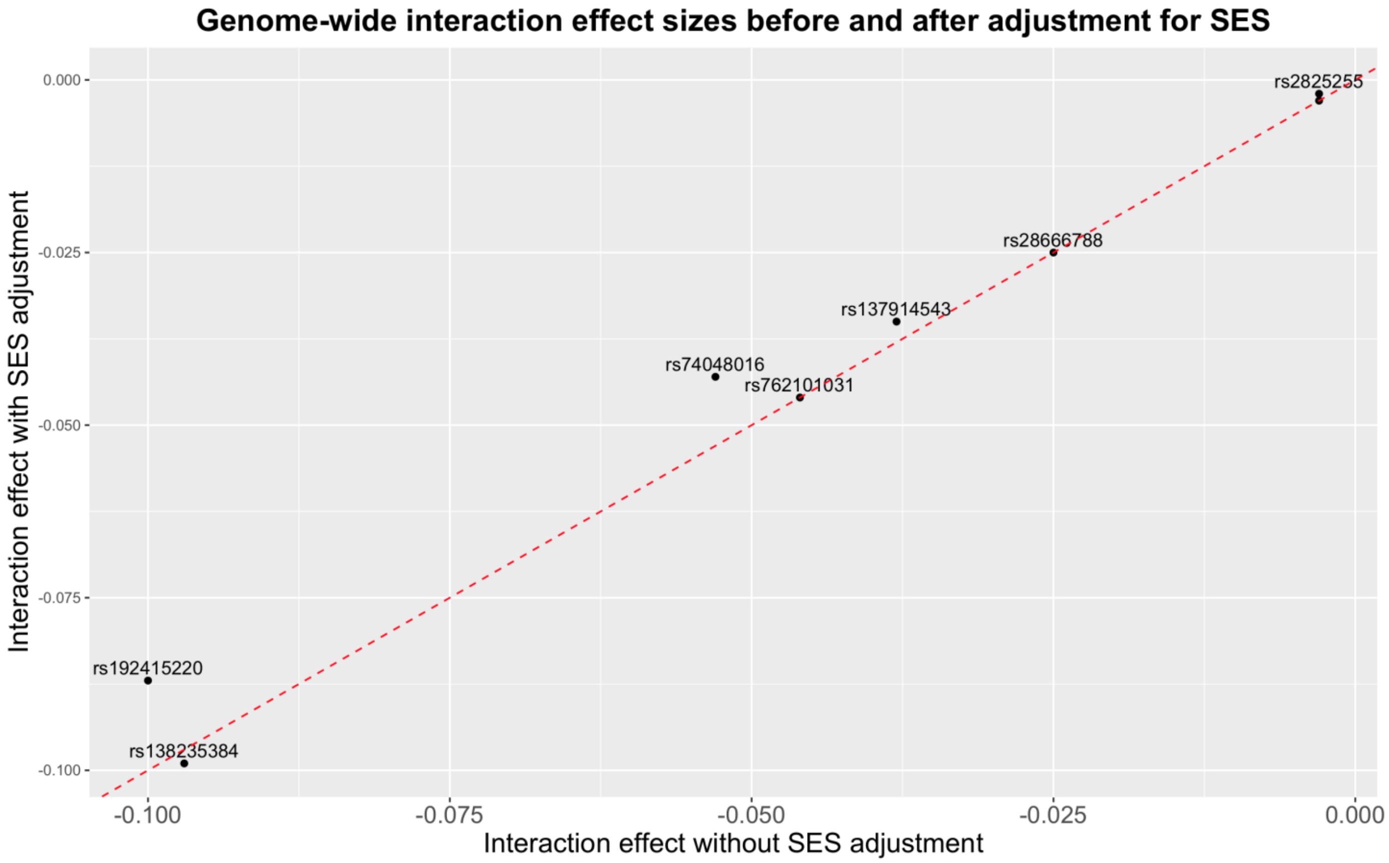

Supplementary figure 8: Interaction effects stratified by education group

(SD = standard deviation, Education group is coded as 0 - Lower vocational qualification or less and 1 – Higher vocational qualification or more)

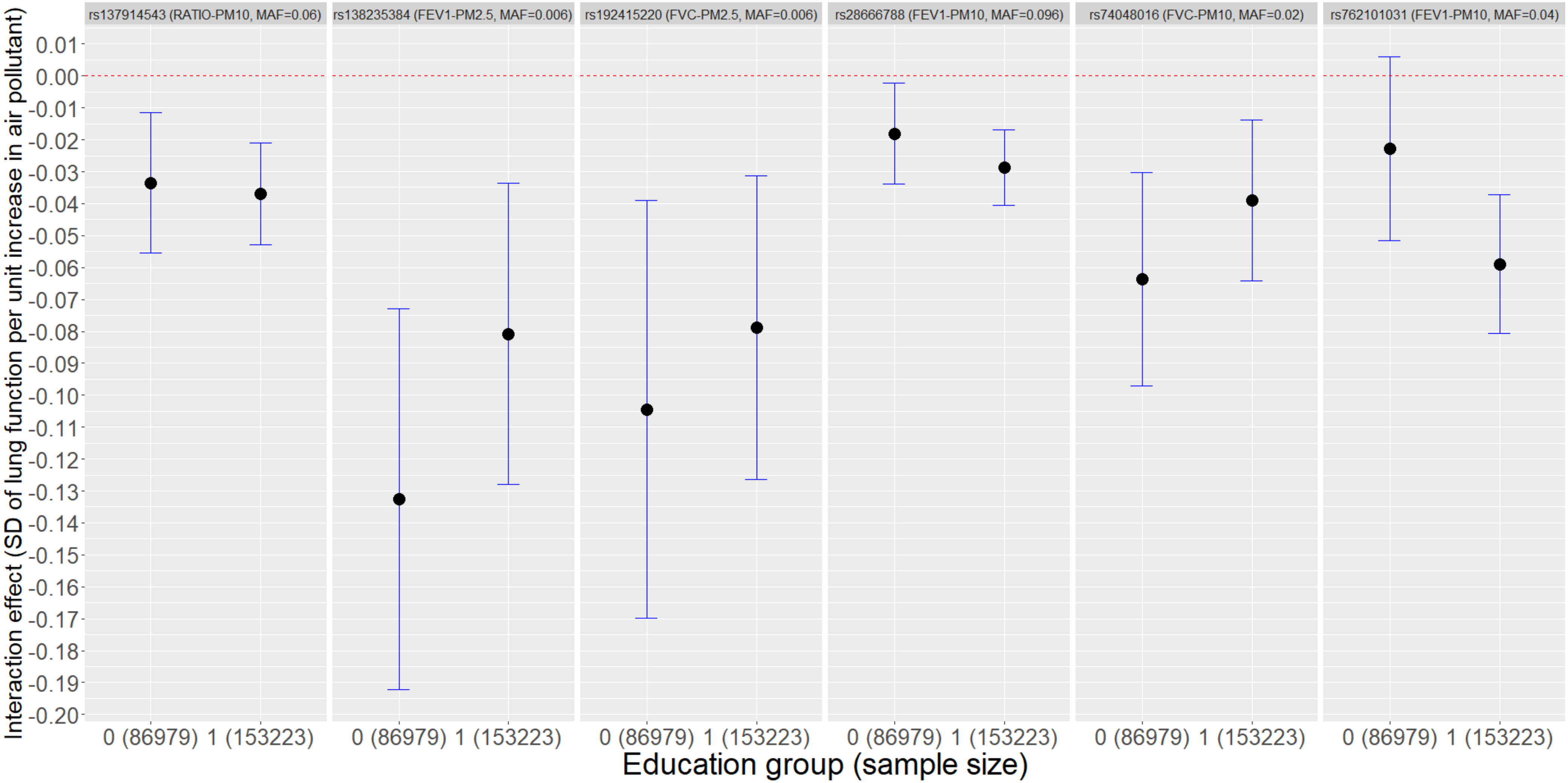

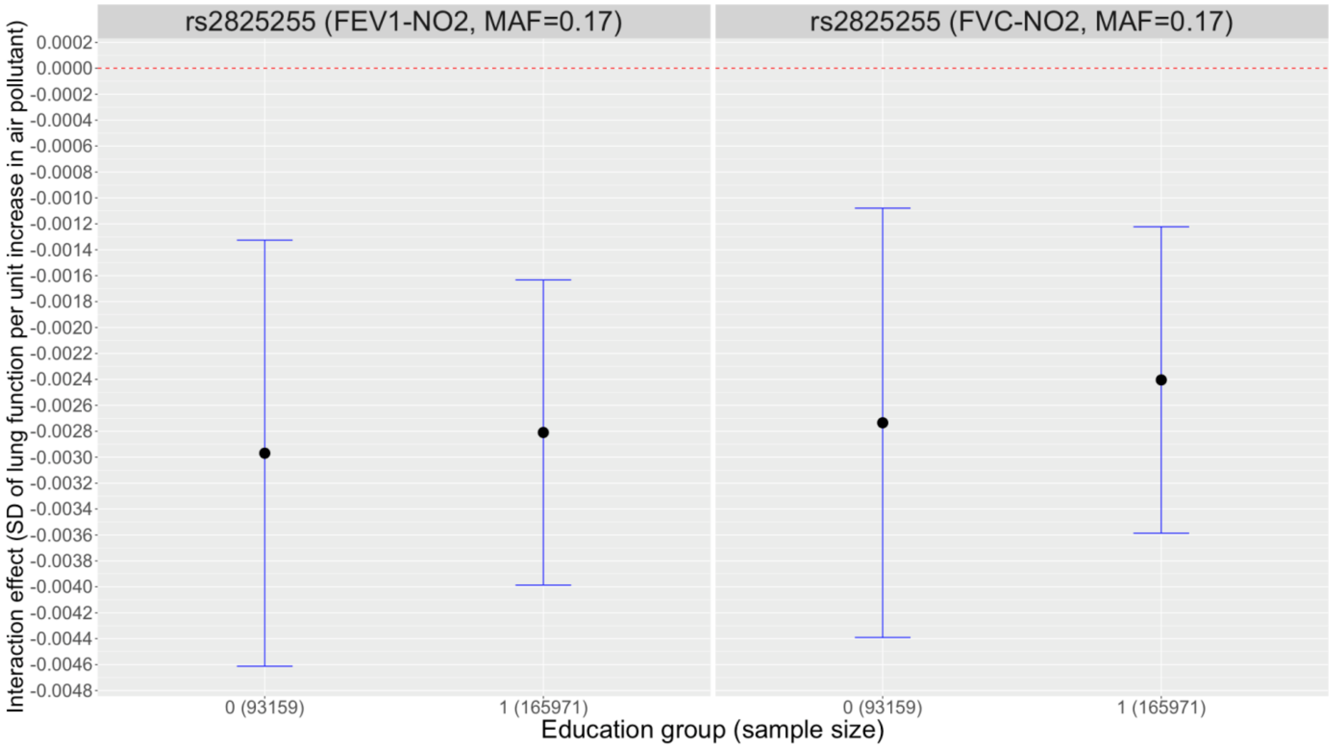

rs2825255 (FEV1-NO2, MAF=0.17)
rs2825255 (FVC-NO2, MAF=0.17)
Interaction effect (SD of lung function per unit increase in air pollutant)
0.0002
0.0000
-0.0002
-0.0004
-0.0006
-0.0008
-0.0010
-0.0012
-0.0014
-0.0016
-0.0018
-0.0020
-0.0022
-0.0024
-0.0026
-0.0028
-0.0030
-0.0032
-0.0034
-0.0036
-0.0038
-0.0040
-0.0042
-0.0044
-0.0046
-0.0048
0 (93159)
1 (165971)
0 (93159)
1 (165971)
Education group (sample size)

Supplementary figure 9: Interaction effects stratified by income group
(SD – Standard Deviation, Income group is coded as 0 - lowest income group to 5 - highest income group)

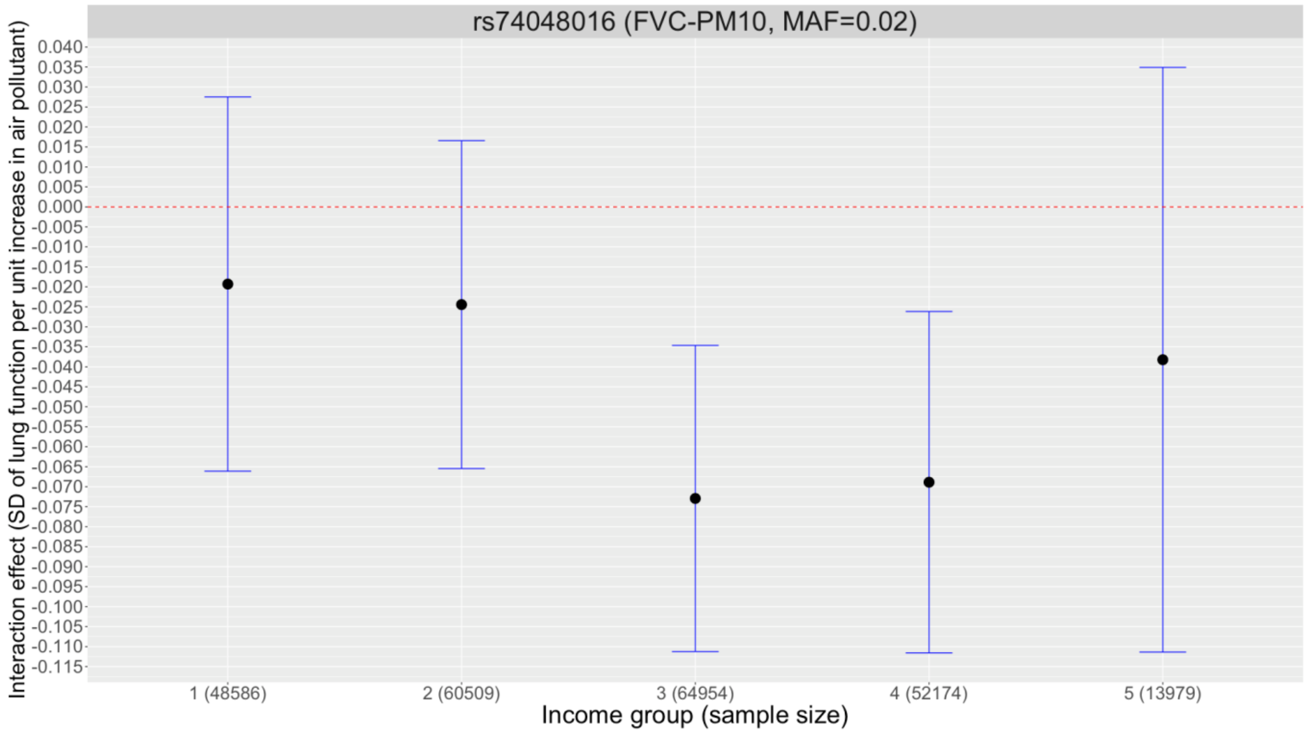

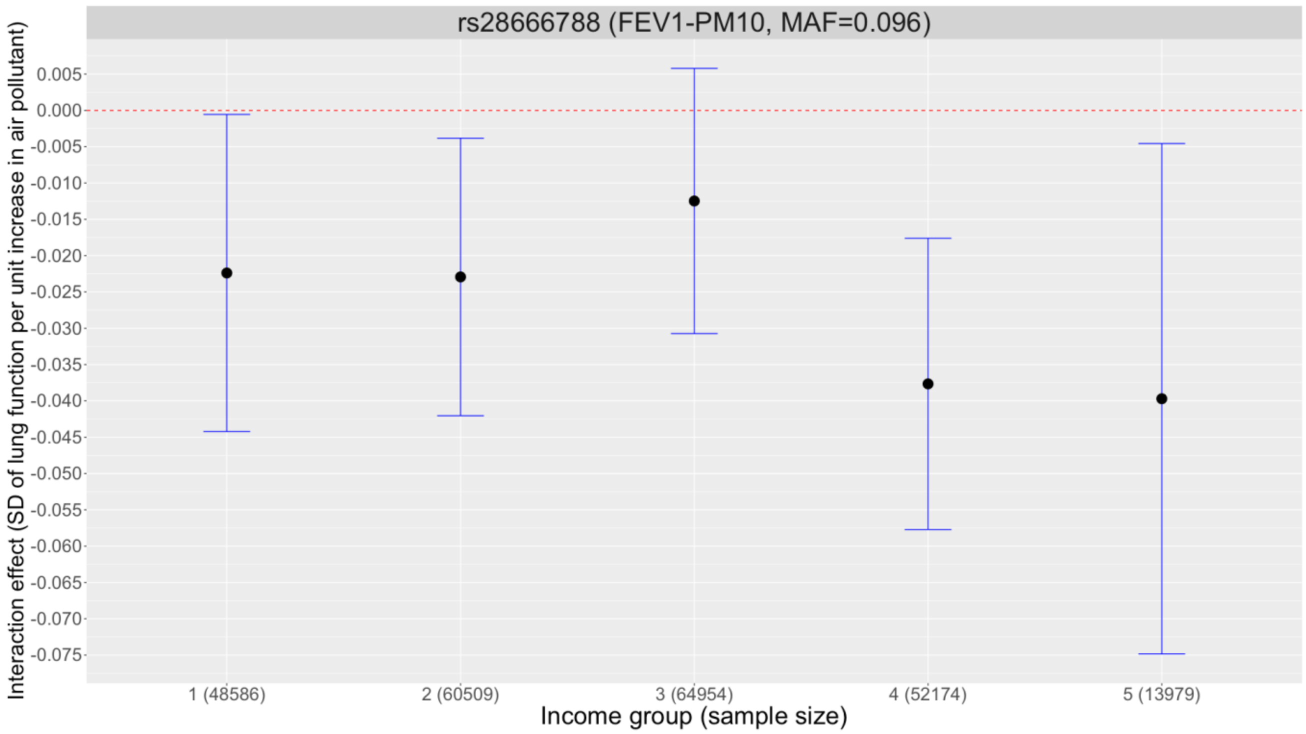

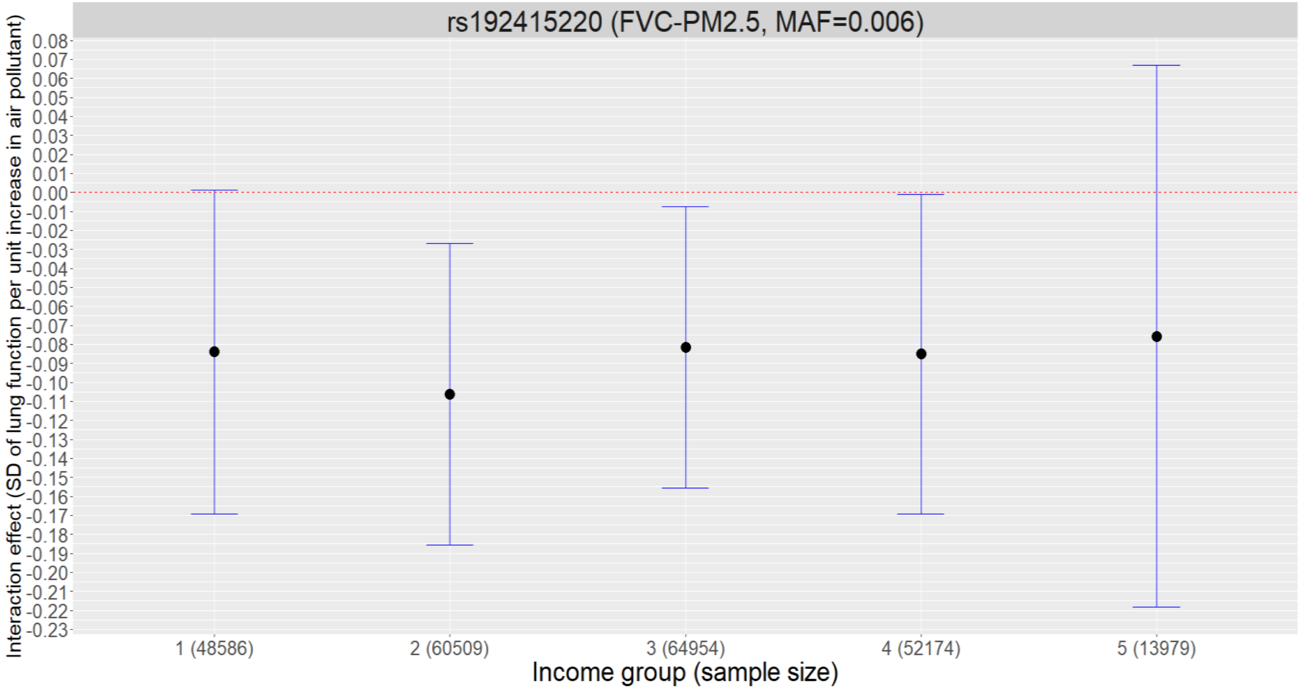
rs192415220 (FVC-PM2.5, MAF=0.006)
Interaction effect (SD of lung function per unit increase in air pollutant)
0.08
0.07
0.06
0.05
0.04
0.03
0.02
0.01
0.00
-0.01
-0.02
-0.03
-0.04
-0.05
-0.06
-0.07
-0.08
-0.09
-0.10
-0.11
-0.12
-0.13
-0.14
-0.15
-0.16
-0.17
-0.18
-0.19
-0.20
-0.21
-0.22
-0.23
1 (48586)
2 (60509)
3 (64954)
4 (52174)
5 (13979)
Income group (sample size)

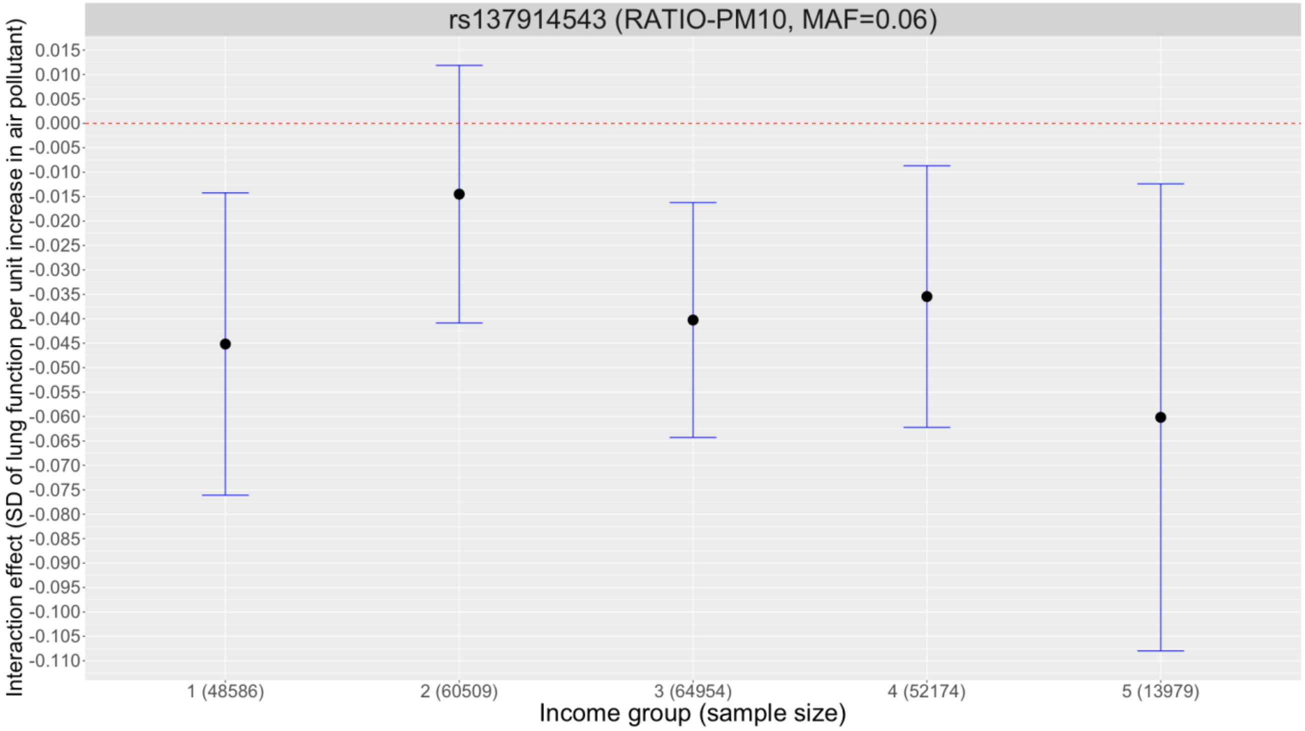
rs137914543 (RATIO-PM10, MAF=0.06)
Interaction effect (SD of lung function per unit increase in air pollutant)
0.015
0.010
0.005
0.000
-0.005
-0.010
-0.015
-0.020
-0.025
-0.030
-0.035
-0.040
-0.045
-0.050
-0.055
-0.060
-0.065
-0.070
-0.075
-0.080
-0.085
-0.090
-0.095
-0.100
-0.105
-0.110
1 (48586)
2 (60509)
3 (64954)
4 (52174)
5 (13979)
Income group (sample size)

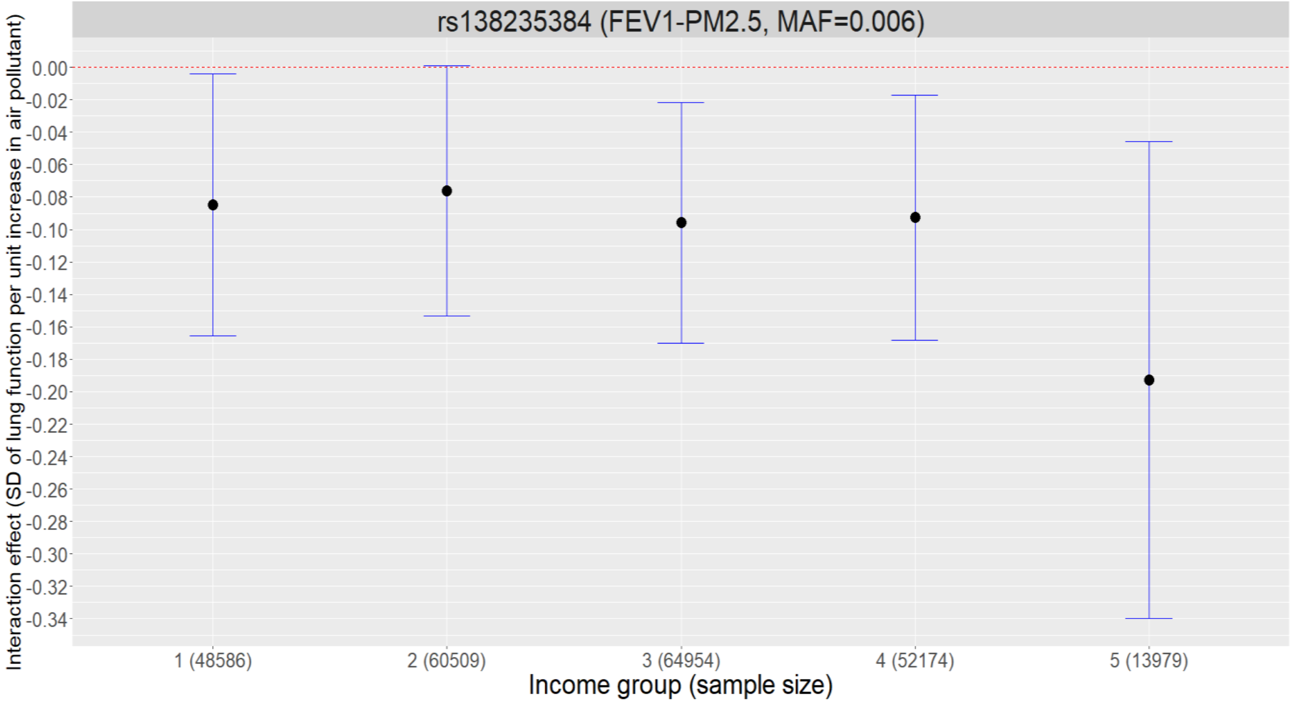
rs138235384 (FEV1-PM2.5, MAF=0.006)
Interaction effect (SD of lung function per unit increase in air pollutant)
0.00
-0.02
-0.04
-0.06
-0.08
-0.10
-0.12
-0.14
-0.16
-0.18
-0.20
-0.22
-0.24
-0.26
-0.28
-0.30
-0.32
-0.34
1 (48586)
2 (60509)
3 (64954)
4 (52174)
5 (13979)
Income group (sample size)

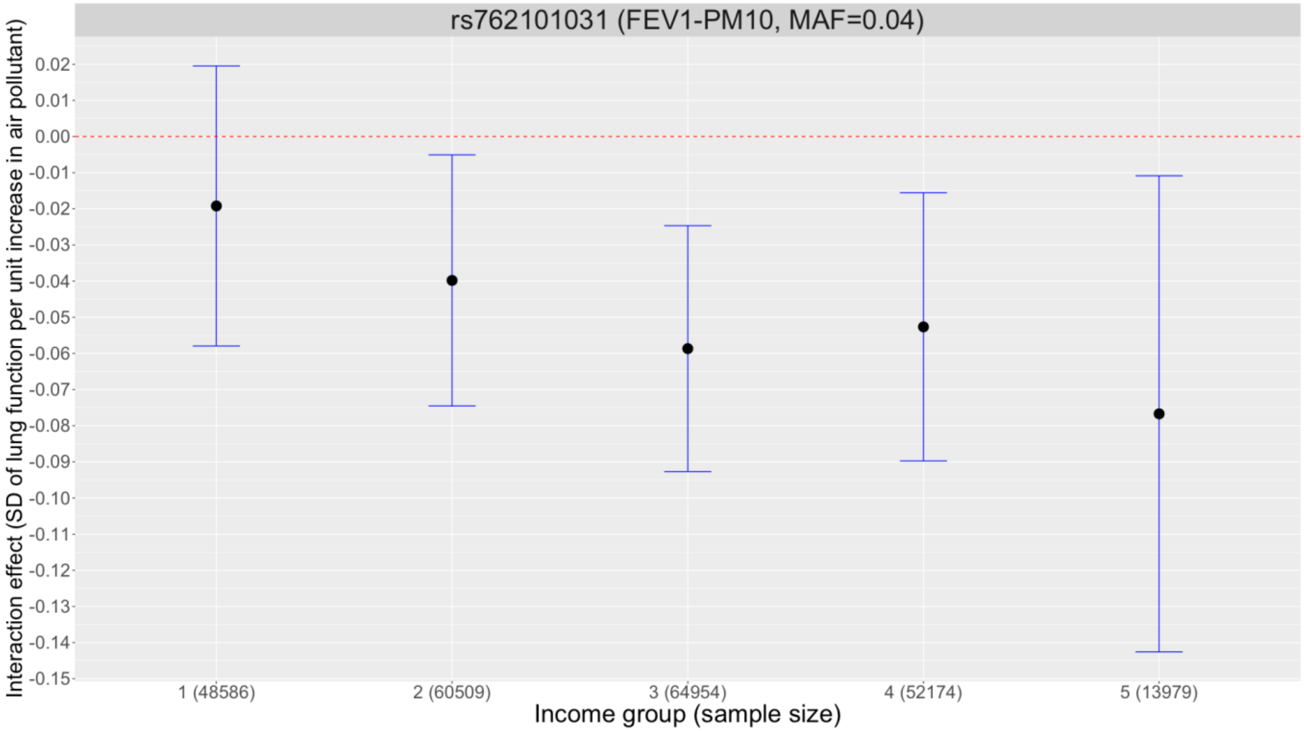
rs762101031 (FEV1-PM10, MAF=0.04)
Interaction effect (SD of lung function per unit increase in air pollutant)
0.02
0.01
0.00
-0.01
-0.02
-0.03
-0.04
-0.05
-0.06
-0.07
-0.08
-0.09
-0.10
-0.11
-0.12
-0.13
-0.14
-0.15
1 (48586)
2 (60509)
3 (64954)
4 (52174)
5 (13979)
Income group (sample size)

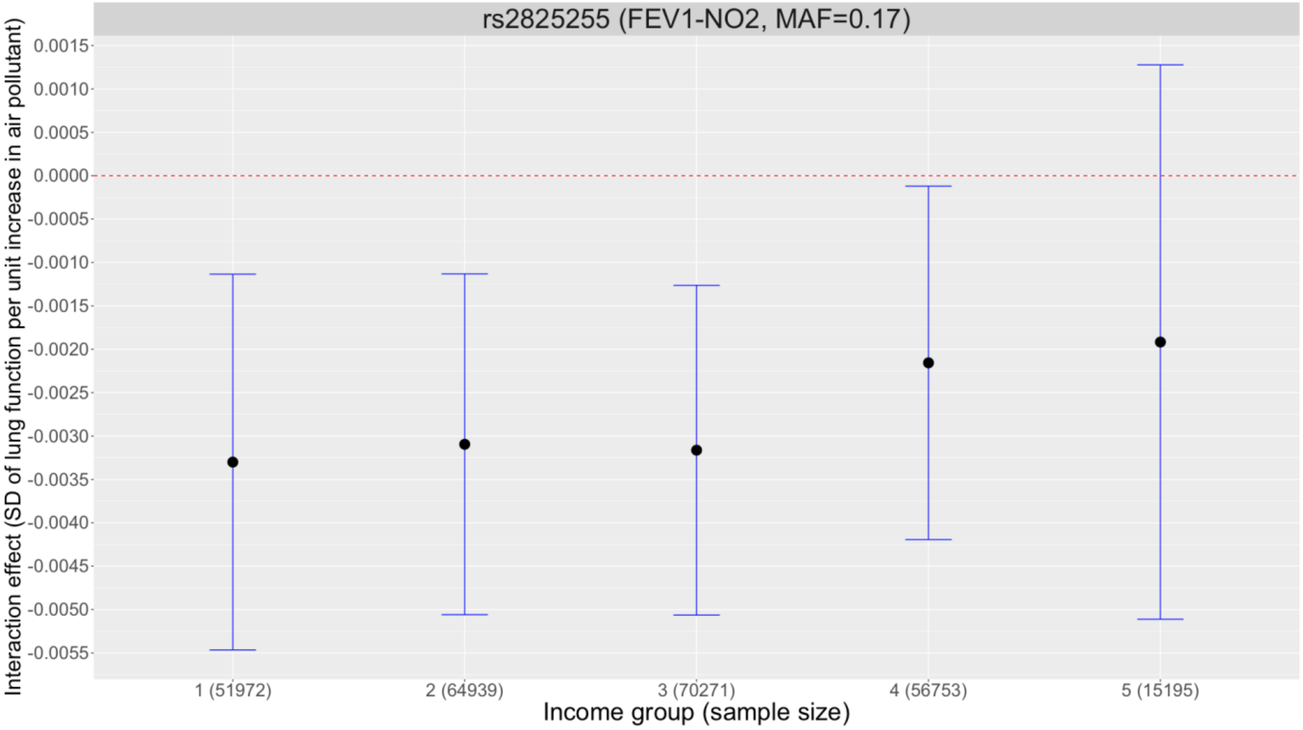
rs2825255 (FEV1-NO2, MAF=0.17)
Interaction effect (SD of lung function per unit increase in air pollutant)
0.0015
0.0010
0.0005
0.0000
-0.0005
-0.0010
-0.0015
-0.0020
-0.0025
-0.0030
-0.0035
-0.0040
-0.0045
-0.0050
-0.0055
1 (51972)
2 (64939)
3 (70271)
4 (56753)
5 (15195)
Income group (sample size)

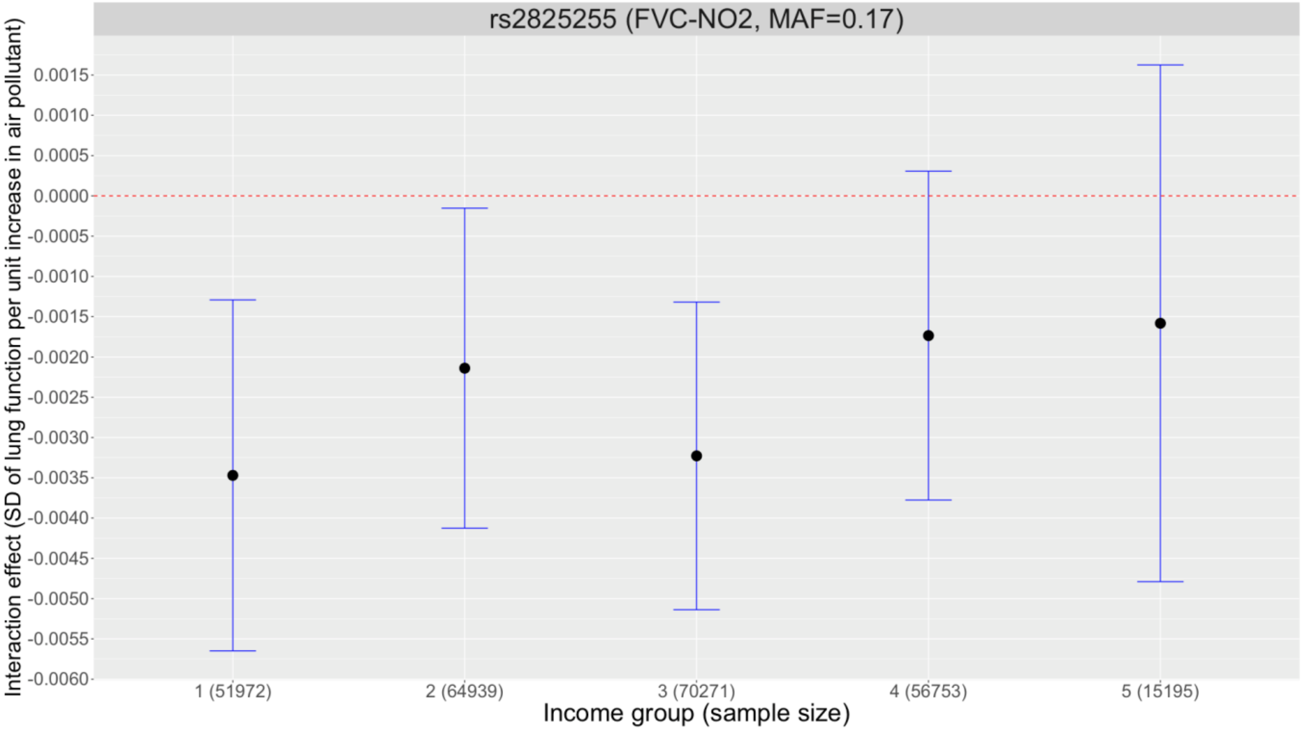
rs2825255 (FVC-NO2, MAF=0.17)
Interaction effect (SD of lung function per unit increase in air pollutant)
0.0015
0.0010
0.0005
0.0000
-0.0005
-0.0010
-0.0015
-0.0020
-0.0025
-0.0030
-0.0035
-0.0040
-0.0045
-0.0050
-0.0055
-0.0060
1 (51972)
2 (64939)
3 (70271)
4 (56753)
5 (15195)
Income group (sample size)
